# Improving Antenatal Syphilis Screening and Treatment Coverage in Public Health Facilities in Kampala, Uganda: A Pre-Implementation and Implementation-Phase Study

**DOI:** 10.64898/2026.10.22.26366192

**Authors:** Annet A. Onzia, Agnes B. Naggirinya, Anna-Beatrice Magoro, Flavia Dhikusooka, Rachel King, Matthew M. Hamill, Yukari C. Manabe, Rosalind Parkes-Ratanshi

## Abstract

**Introduction:** Congenital syphilis (CS) remains a major cause of preventable adverse pregnancy outcomes globally, driven by poor antenatal care (ANC) quality, health-system barriers, and untreated maternal infection. In Uganda, syphilis screening and treatment coverage remains below national targets. We evaluated service-delivery impact of a multi-component syphilis testing and treatment intervention implemented across four public health facilities in Kampala, Uganda.

**Methods:** Guided by the Consolidated Framework for Implementation Research (CFIR), the intervention introduced same-day dual HIV/syphilis antibody testing, on-site treatment, and partner notification at three ANC facilities with women with serological evidence of syphilis or unknown syphilis status and infant follow-up at a tertiary referral hospital. Facility register data were compared between a pre-implementation period (January-December 2020) and the implementation period (June 2021-July 2022) using a modified Poisson regression model adjusting for site.

**Results:** Syphilis screening among women attending their first ANC visit increased from 55.2% to 88.3% (+33.6%; 95% CI 21.0-46.3; p<0.001); same-day treatment among those testing positive increased from 74% to 100% (difference 26.1; 95% CI 22.4-29.9; p<0.001). Partner testing increased nearly five-fold, from 11.8% to 62% (difference 51.9; 95% CI 46.8-57.0; p<0.001); 23.6% of tested partners were positive. Among the high-risk women, 62% tested positive for syphilis, nearly fifteen-fold higher than the general ANC population, and up to 46% of their infants met criteria for confirmed or possible CS; 41% of whom completed intravenous treatment.

**Conclusions:** A same-day rapid test-and-treat intervention substantially improved maternal syphilis screening and treatment coverage, even during the COVID-19 pandemic, but partner and infant treatment completion remained major gaps in the care cascade. Structural interventions, including integrated maternal-infant care pathways and targeting of high-risk mothers along with their male partners, are needed to close these gaps and advance progress toward the elimination of mother-to-child transmission of syphilis.

**What is already known on this topic:** Congenital syphilis (CS) is a major, preventable cause of stillbirth, neonatal death and adverse pregnancy outcomes, driven by missed or late screening, delayed treatment and untreated maternal infection. In Uganda, antenatal syphilis screening and treatment fall below national targets, and it is unclear whether health-system interventions can close these gaps in routine public facilities.

**What this study adds:** A multi-component intervention (same-day dual HIV/syphilis testing, on-site treatment and partner notification) substantially increased first-visit screening in Kampala public facilities, and all women who tested positive were treated the same day. Partner testing rose several-fold, and many tested partners were positive, revealing a large untreated reservoir. High-risk women had much higher syphilis positivity than the general antenatal population. Nearly half of their infants met criteria for confirmed or possible CS, yet fewer than half of these infants completed intravenous treatment.

**How this study might affect research, practice or policy:** Same-day test-and-treat is feasible in routine ANC and could be scaled nationally, with targeting of high-risk women and their male partners. Partner and infant treatment completion are the weakest points in the care cascade, so integrated maternal-infant care pathways are needed to advance elimination of mother-to-child transmission. Future research should test strategies to improve partner treatment and infant follow-up and assess the effect on CS incidence and cost-effectiveness.

## Introduction

Syphilis remains a significant public health issue globally (1). In addition to increasing the risk of HIV transmission (2), untreated maternal syphilis increases the risk of adverse pregnancy outcomes, with 50-80% of cases resulting in stillbirth, abortion, prematurity, or newborn congenital syphilis (CS) (3). CS occurs when the bacterium *Treponema pallidum* passes through the placenta from mother to fetus or through contact with infectious syphilitic lesions during delivery (4).

CS is the second leading cause of preventable stillbirths globally after malaria (5) and its global burden remains substantial. In 2022, the World Health Organization (WHO) estimated over 700,000 cases worldwide, causing ∼390,000 adverse birth outcomes (150,000 early fetal deaths/stillbirths, 70,000 neonatal deaths, and 55,000 preterm or low-weight births), with over 62% concentrated in Africa (6). Although effective, inexpensive treatment exists, CS persists due to poor quality antenatal care (ANC), health system barriers, Benzathine penicillin (BPG) stockouts, and reinfection from untreated partners (7). Estimates of CS in individual African countries vary widely (8, 9). Our STOP (Syphilis Treatment of Partners) trial found a 3.5% maternal syphilis prevalence; untreated male partners were significantly associated with adverse birth outcomes (OR 2.75, 95% CI 2.36-3.21, p<0.001) (10).

In 2016, WHO established global targets for the elimination of mother-to-child transmission (EMTCT) of syphilis, with the ultimate goal of reducing congenital syphilis to ≤50 cases per 100,000 live births in at least 80% of countries by 2030 (11). In response to this, free same-day dual HIV/syphilis screening and immediate treatment in ANC were incorporated into Uganda’s national EMTCT guidelines in 2017. Although ANC attendance in Uganda is high, important gaps remain in timely initiation and continuity of care. Only approximately 37% of women initiate ANC in the first or second trimester, and 68% complete at least four ANC visits (12). These gaps, together with workforce shortages, long waiting times, and disruptions in the supply chain limit opportunities to achieve the WHO target of ≥95% syphilis testing and treatment among pregnant women attending ANC (9, 13).

Despite these gaps, few studies in Uganda have rigorously evaluated whether a multi-component, implementation research framework-guided service-delivery intervention can close the gap between national targets and actual screening and treatment coverage. To address this, the Kampala Syphilis Screening and Treatment (KSST) project was implemented from June 2021 to July 2022 at high-volume public health facilities in Kampala District, Uganda. This manuscript reports quantitative service-delivery outcomes from KSST, comparing pre-implementation and implementation-period coverage across the mother-partner-infant care cascade.

## Methods

### Project setting

KSST project was conducted at four public health facilities providing antenatal, delivery, and postnatal care in Kampala District, Uganda: Kisenyi, Kawaala, and Kiswa Health Centers IV, and a tertiary facility, Kawempe National Referral Women’s Hospital (**Supplemental File 1**).

### Project population

At the three lower-level health facilities, the project enrolled all pregnant women attending their first ANC visit (<27 weeks’ gestation), and the male partners of women who tested positive, by rapid diagnostic test (RDT), for treponemal antibodies. At Kawempe National Referral Hospital, which has a dedicated maternal wing, we enrolled women considered ‘high-risk’, defined as those who had either never been tested for syphilis during pregnancy before 36 weeks of gestation, or those who tested positive in the third trimester of pregnancy, during labor/delivery, or postnatally.

### Pre-project needs assessment

#### Kisenyi, Kawaala and Kiswa Health facilities

Guided by the Consolidated Framework for Implementation Research (CFIR) (14), we designed and implemented an intervention to improve syphilis testing and treatment uptake, systematically assessing multi-level barriers and facilitators.

Before implementation, data were extracted on syphilis screening and treatment over the prior 12 months at the three facilities to assess needs and shortfalls (15). Project nurses, including a monitoring and evaluation specialist, reviewed routine data collected between January and December 2020. This included the number of women attending their first ANC, those screened, testing positive, and treated for syphilis, proportion notifying partners, partners tested and treated, and infants tested and treated for CS. The findings identified gaps in service provision, informing implementation strategies to enhance facility readiness, improve fidelity to national guidelines, and support effective service delivery (**Table 1**).

**Table 1:**
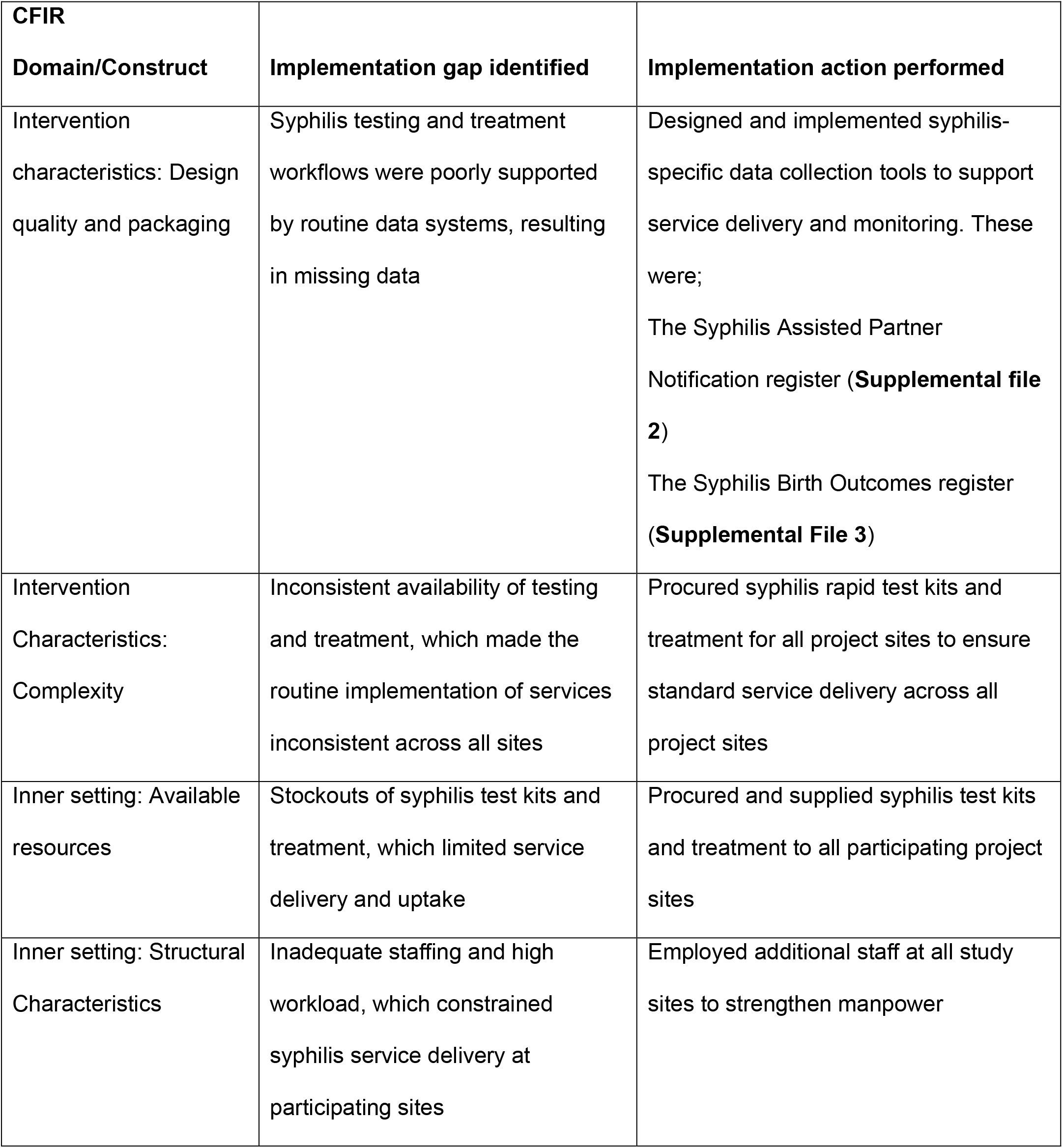
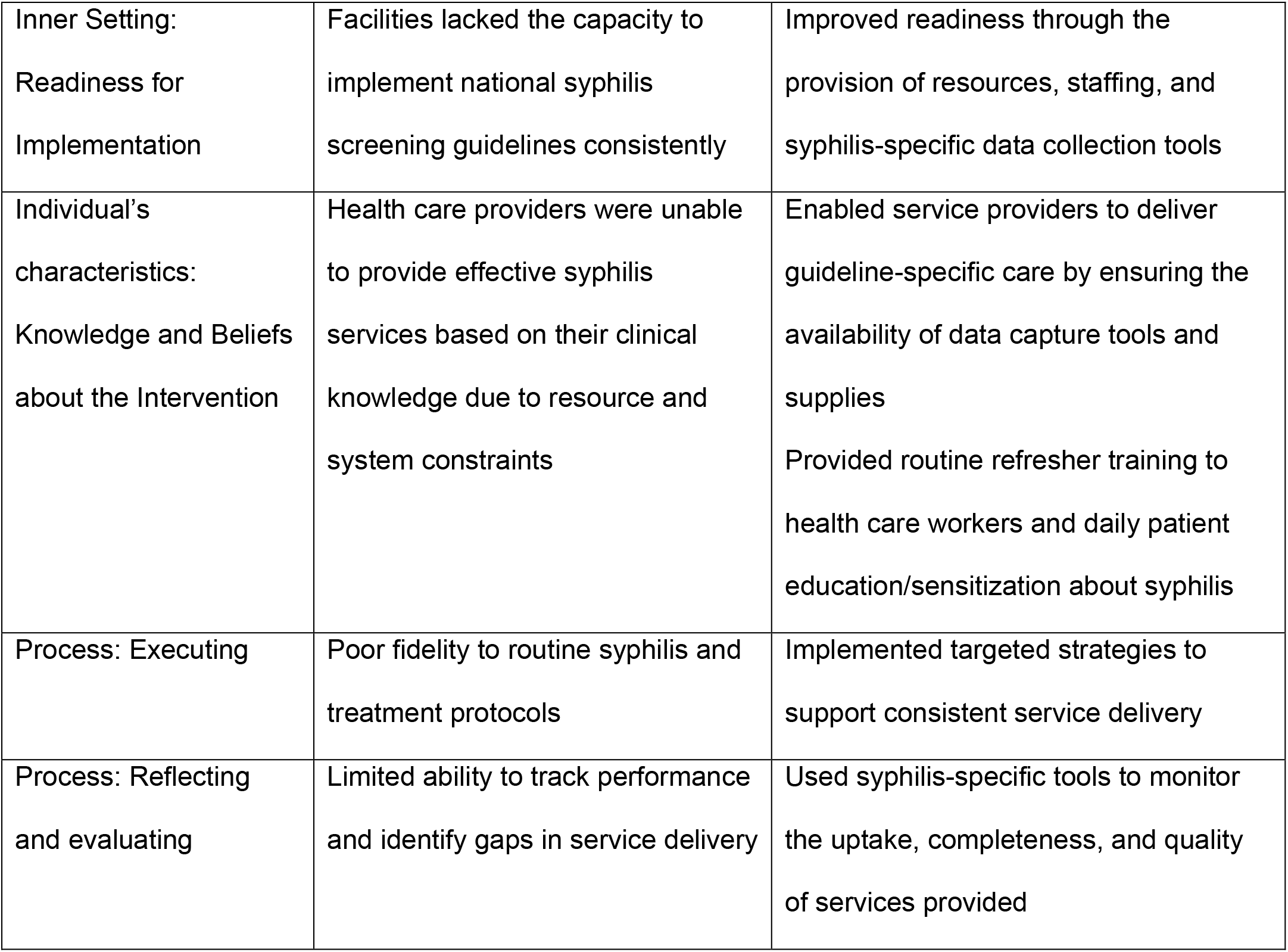
Project needs assessment guided by the Consolidated Framework for Implementation Research.

| CFIR<br>Domain/Construct | Implementation gap identified | Implementation action performed |
| --- | --- | --- |
| Intervention<br>characteristics: Design<br>quality and packaging | Syphilis testing and treatment<br>workflows were poorly supported<br>by routine data systems, resulting<br>in missing data | Designed and implemented syphilis-specific data collection tools to support<br>service delivery and monitoring. These<br>were;<br>The Syphilis Assisted Partner<br>Notification register (**Supplemental file**<br>**2**)<br>The Syphilis Birth Outcomes register<br>(**Supplemental File 3**) |
| Intervention<br>Characteristics:<br>Complexity | Inconsistent availability of testing<br>and treatment, which made the<br>routine implementation of services<br>inconsistent across all sites | Procured syphilis rapid test kits and<br>treatment for all project sites to ensure<br>standard service delivery across all<br>project sites |
| Inner setting: Available<br>resources | Stockouts of syphilis test kits and<br>treatment, which limited service<br>delivery and uptake | Procured and supplied syphilis test kits<br>and treatment to all participating project<br>sites |
| Inner setting: Structural<br>Characteristics | Inadequate staffing and high<br>workload, which constrained<br>syphilis service delivery at<br>participating sites | Employed additional staff at all study<br>sites to strengthen manpower |
| Inner Setting:<br>Readiness for<br>Implementation | Facilities lacked the capacity to<br>implement national syphilis<br>screening guidelines consistently | Improved readiness through the<br>provision of resources, staffing, and<br>syphilis-specific data collection tools |
| Individual's<br>characteristics:<br>Knowledge and Beliefs<br>about the Intervention | Health care providers were unable<br>to provide effective syphilis<br>services based on their clinical<br>knowledge due to resource and<br>system constraints | Enabled service providers to deliver<br>guideline-specific care by ensuring the<br>availability of data capture tools and<br>supplies<br>Provided routine refresher training to<br>health care workers and daily patient<br>education/sensitization about syphilis |
| Process: Executing | Poor fidelity to routine syphilis and<br>treatment protocols | Implemented targeted strategies to<br>support consistent service delivery |
| Process: Reflecting<br>and evaluating | Limited ability to track performance<br>and identify gaps in service delivery | Used syphilis-specific tools to monitor<br>the uptake, completeness, and quality<br>of services provided |

The needs assessment informed project targets, including syphilis screening by RDT for 95% of women at first ANC visit; stage-appropriate syphilis treatment for 90% of those testing positive; partner-notification counselling for 90% of treponemal antibody-positive women; and testing and treatment for 60% of their partners. The assessment also identified a gap in documentation of outcomes, leading to the development of an electronic Syphilis Assisted Partner Notification (APN) register (**supplemental file 2**), based on the existing APN services for HIV.

#### Kawempe National Referral Hospital

Assigned project staff reviewed routine data collected from January to December 2020, including the number of women whose pregnancies were considered at high risk of CS, the numbers screened, testing positive, and treated for syphilis; the proportion notifying partners; the proportion of partners tested and treated; and the number of infants followed up, tested, diagnosed, and treated for CS. This led to the development of the Syphilis Birth Outcomes Register (SBOR) (**supplemental file 3**), which was used to collect data at the facility during project implementation.

### Project Implementation

#### Kisenyi, Kawaala and Kiswa Health facilities

Between June 2021 and July 2022, data were collected at Kisenyi, Kawaala, and Kiswa Health Center IV using the Syphilis APN registers by project staff and facility-based midwives. This period coincided with the COVID-19 lockdown measures in Uganda which challenged health services. Project staff conducted sensitization sessions on syphilis and its management. Women who tested treponemal antibody rapid test-positive using the SD Bioline™ HIV/syphilis Duo test (Abbott, Illinois, Abbot Point of Care, Princeton, NJ, USA) initiated same-day treatment with BPG, were counseled to complete all doses, notify their partners, with couple testing and treatment done according to local guidelines (16). Partners were offered a single dose BPG following a positive rapid antibody test. Due to funding constraints, confirmatory syphilis testing was not done at these lower-level facilities. Monthly register reviews at each site tracked service-delivery outcomes, including the proportion of women and partners screened, tested, and treated (**Supplemental File 4**).

#### Kawempe National Referral Hospital

Women whose pregnancies were considered at risk of CS were referred to Kawempe Women’s Hospital, identified either directly at the hospital or through referral from the three lower-level sites. Kawempe was selected for its tertiary capacity and specialist gynecologists and pediatricians able to manage complex CS cases. Data were collected using SBOR. A Serologic testing reactive maternal rapid syphilis treponemal antibody test, SD Bioline™ HIV/syphilis Duo test (Abbott, Illinois, Abbot Point of Care, Princeton, NJ, USA) result prompted confirmatory testing using the lipoidal rapid plasma reagin (RPR) (BD Macro-Vue RPR card test, Becton Dickinson, Sparks, Maryland, USA), confirmed with the specific *Treponema pallidum* hemagglutination test (TPHA) assay (MonlabTest® Omega Diagnostics, Scotland, United Kingdom). A positive syphilis test comprised a reactive RPR and TPHA result. RPR titer by serial dilution was performed on sera of those who tested positive by RPR and TPHA. High titer was defined as an RPR titer of ≥1:8, as described in previous studies (8, 17, 18). Partners of women with a positive antibody test were offered testing, and same-day treatment was initiated for those with a positive antibody test result.

Pregnancy outcomes were tracked, and babies born to women with RPR titers ≥1:8 were followed up after delivery and examined by a pediatrician, with serum RPR results typically available within 1-3 days. Same-day maternal and infant serum (or cord blood) RPR titers were compared, and CS was classified as adapted for low-resource settings per Centers for Disease Control (CDC) guidelines (19). This involved relying on clinical assessment, maternal syphilis history and treatment status, and any available laboratory investigations, as confirmatory investigations recommended in the CDC guidelines, like cerebrospinal fluid examination and long-bone radiographs, were not routinely available. Parents were offered treatment for their infant with a 10-day course of intravenous aqueous crystalline Penicillin G, according to local guidelines (16). Follow-up phone calls were made to women with reactive treponemal RDTs to ascertain pregnancy outcomes after delivery (**Supplemental File 4**).

### Data analysis

Data were manually extracted from the registers, with key outcomes tallied as daily counts, and proportions calculated. Pre-implementation and during-implementation outcomes were compared using proportions and differences. Risk differences and 95% confidence intervals (CIs) were obtained from a modified Poisson regression model with robust standard errors, adjusting for site. Study findings are reported according to the Standards for Reporting Implementation Studies (StaRI) Checklist (**Supplemental File 5**).

### Patient and Public Involvement Statement

Selected members of the public were involved in the design and conduct of this study. Policymakers, service providers, local leaders and women attending ANC reviewed the registers to ensure the questions were clear, culturally appropriate, and relevant to health concerns which facilitated participant engagement. Findings were disseminated to the Ministry of Health STI Control Unit to promote awareness and inform future interventions.

### Ethical approval

The Infectious Diseases Institute Research Ethics Committee (IDI REC 008/2022) and the Uganda National Council for Science and Technology (HS2436ES) approved the study. Informed consent was waived for this implementation phase, in line with standard Ministry of Health procedures for routine service-delivery data collected as part of standard of care.

## Results

### Kisenyi, Kawaala and Kiswa health facilities

#### Needs assessment

Between January and December 2020 (pre-implementation period), 33,831 pregnant women attended their first ANC visit across Kisenyi, Kawaala, and Kiswa health centers. Of these, 55.2% (n=18,684) were screened for syphilis using RDT, 3.9% (n=740) tested positive, and 74% (n=547) initiated same-day treatment (**Figure 1**). Women initiated on treatment were counselled to return for the second and third doses, administered one week apart over the subsequent two weeks. Those who were unable to return to the study facility were provided with their subsequent doses and counseled to complete treatment at the nearest health facility to them. Of women testing positive, 11.8% (n=87) returned with partners for testing; of these, 44.8% (n=39) tested positive by RDT, and 69.2% (n=27) initiated same-day intramuscular BPG per local guidelines (**Figure 2**).

**Figure 1:**
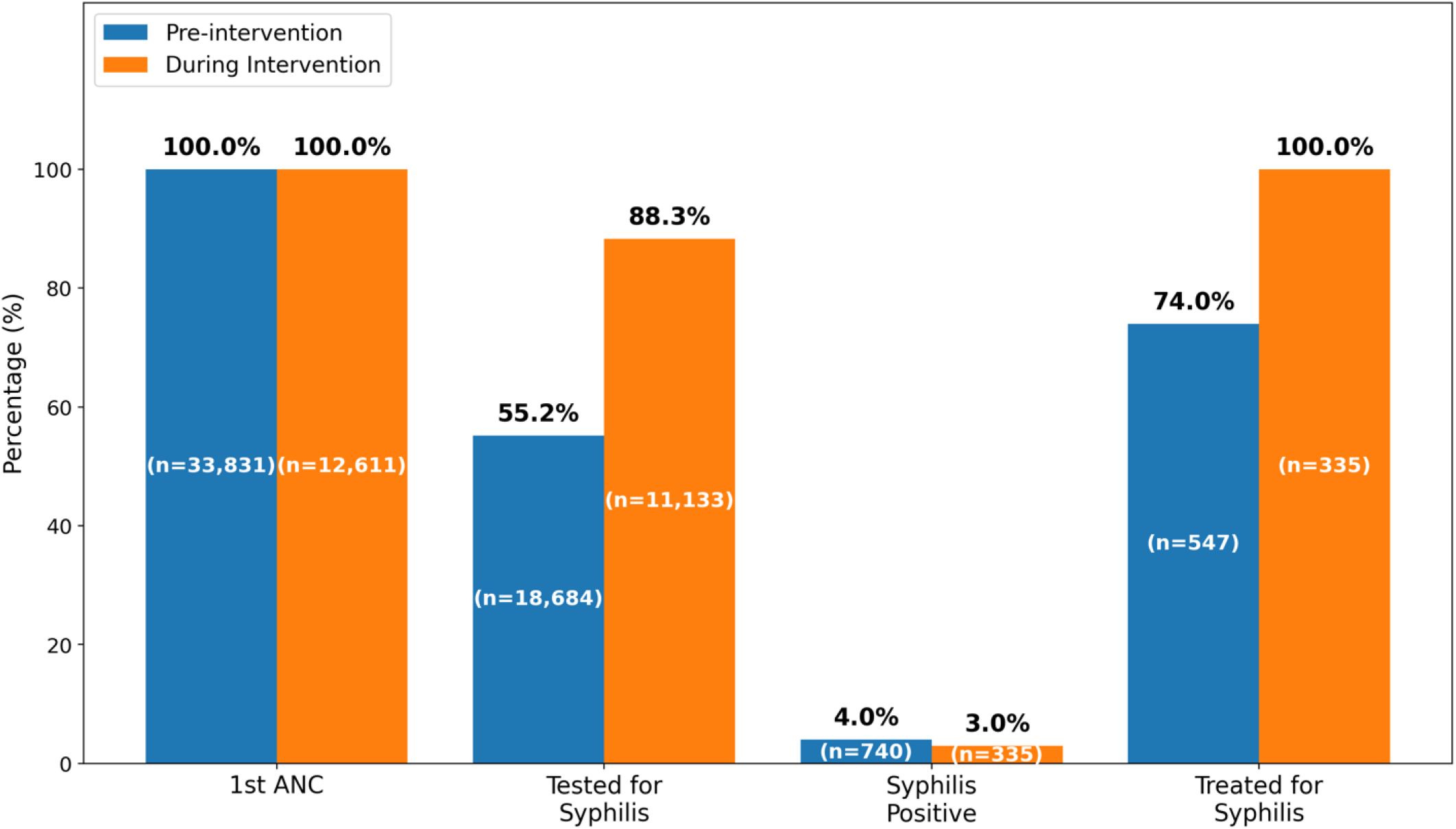
Pregnant women who received syphilis services before and during the intervention, at Kisenyi, Kawaala, and Kiswa health facilities, Kampala, Uganda between the pre-implementation period (January-December 2020) and the implementation period (June 2021-July 2022).

**Figure 2:**
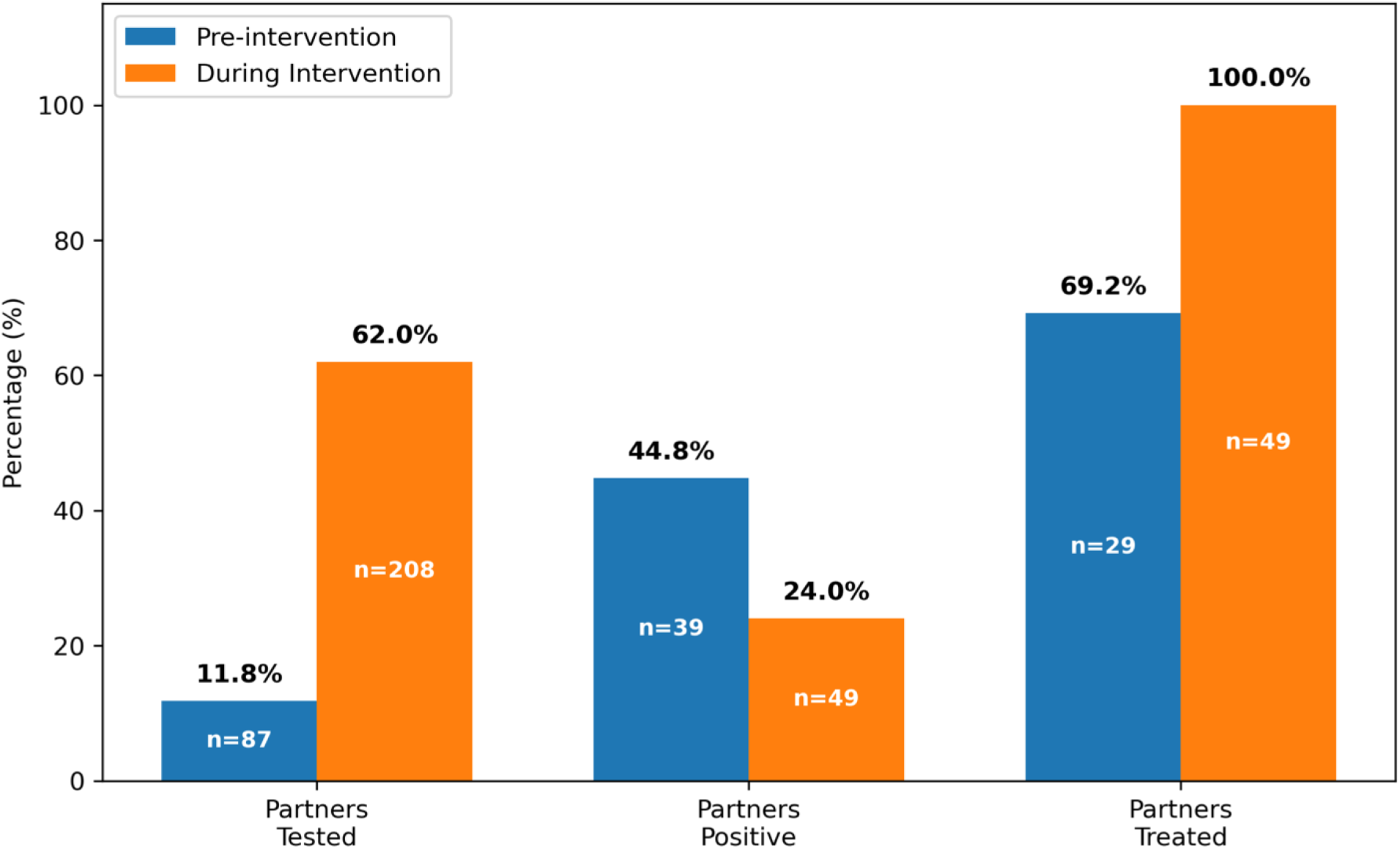
Partners of women who received syphilis services before and during the intervention at Kisenyi, Kawaala, and Kiswa health facilities, Kampala, Uganda, between the pre-implementation period (January-December 2020) and the implementation period (June 2021-July 2022).

#### Project Implementation

Between June 2021 and July 2022, all 12,611 women attending their first ANC visit at the three clinics were sensitized on syphilis testing. Of these, 88.3% (n=11,133) were tested, a significant increase (+33.63; 95% CI 20.99-46.27; p<0.001) compared with pre-KSST implementation (**Table 2**). Among those tested, 3.0% (n=335) were positive, and all initiated same-day treatment, a significant increase (+26.11; 95% CI 22.37-29.86; p<0.001) compared with pre-implementation (**Figure 1**). All 335 women were counselled on partner notification; 62% (n=208) of partners received testing, a significant difference (+51.91; 95% CI 46.78-57.04; p<0.001) compared with pre-KSST. Among partners tested, 23.6% (n=49) were positive by RDT, and all initiated same-day treatment, a significant increase from baseline (difference 29.60; 95% CI 19.95-39.25; p<0.001) (**Figure 2**).

**Table 2:**
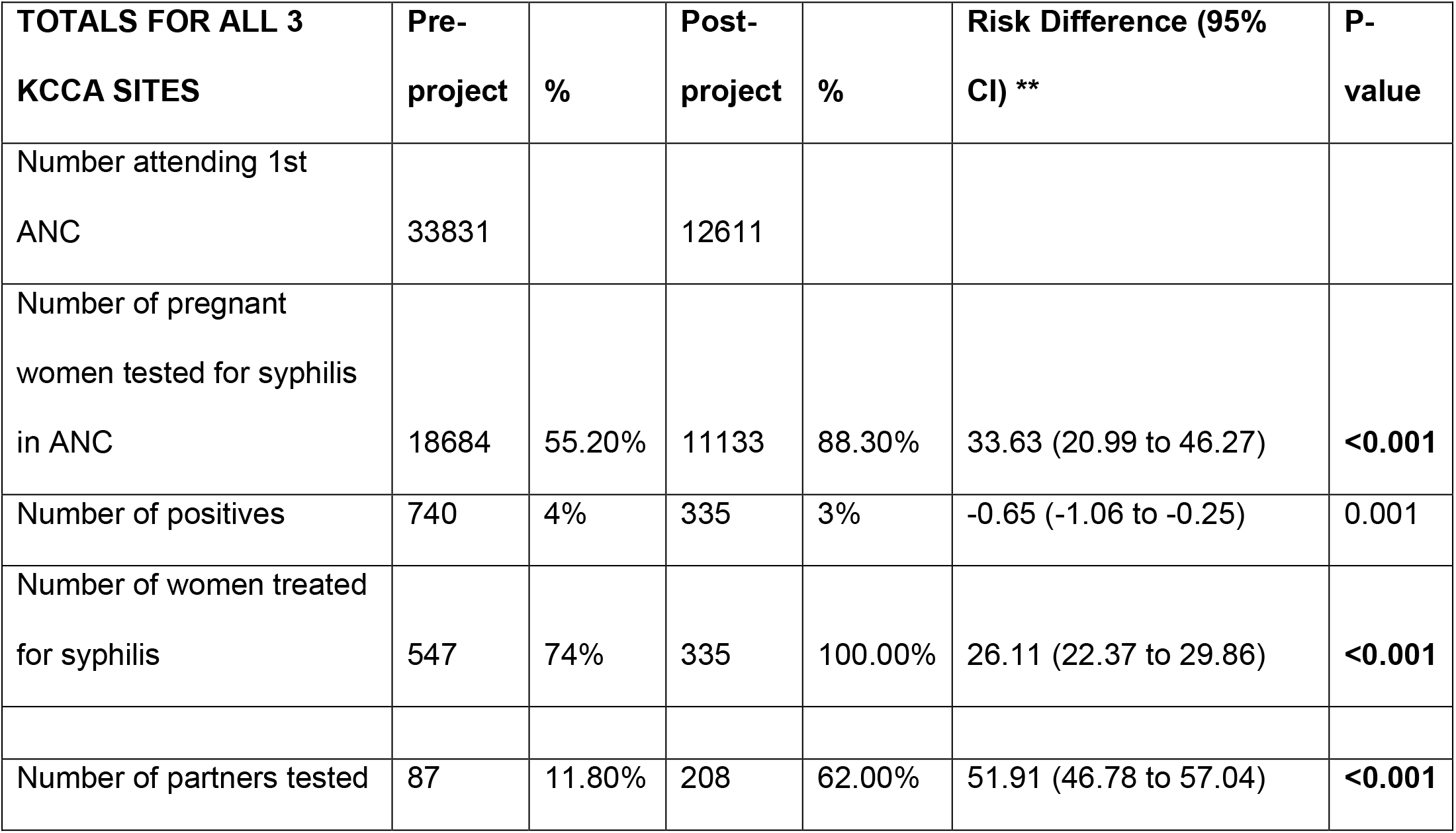
Modified Poisson Regression model for pre-intervention and during-intervention activities among women and partners receiving Antenatal care syphilis services in public health facilities in Kampala, Uganda.

### Kawempe National Referral Hospital

#### Needs Assessment

The project team identified that no dedicated, systematic follow-up existed for women with pregnancies considered at high risk of CS, nor for their infants. All pregnant women were screened for syphilis according to standard practice, irrespective of risk status, with no differentiation in follow-up procedures based on risk level. This led to the development of the SBOR to document pregnancy and infant outcomes and facilitate continuity of care for high-risk mother–infant pairs.

#### During project implementation

125 high-risk women were identified; 62% (n=78) tested positive by SD Bioline. All were initiated on same-day treatment as syphilis of unknown duration, according to local guidelines, with 55% (n=43) receiving all three doses of BPG administered one week apart at the facility (20). However, we were unable to ascertain complete BPG treatment for the remainder. Further testing of the treponemal antibody-positive samples revealed that 41% (n=32) had a reactive RPR test result, with the majority (90.6%, n=29/32) having titers of ≥ 1:8. Of the 78 high-risk women who tested positive by SD Bioline, 69% (n=54) of partners were tested, and 38.9% (n=21) tested positive. All were initiated on same-day treatment with a single dose of BPG.

### Infant outcomes

Among babies born to women with a positive RPR test result, 86.5% (n=26/32) were followed up, either by physical examination after birth or by phone call within one month; six could not return for physical follow-up. On examination, no clinical characteristics of CS were noted, with two babies undergoing treatment for neonatal sepsis. All 26 babies examined were tested for syphilis; 46% (n=12) had a reactive RPR test (titers ranged from 1:4 to 1:64). Of these, three (11.5%) babies (including those being managed for neonatal sepsis) had RPR titers at least fourfold higher than their mothers’ (same-day maternal vs. infant titers; 1:8 vs. 1:32, 1:8 vs. 1:64, and 1:16 vs. 1:64). These three were subsequently managed as confirmed CS as guided by the CDC guidelines (19). The remaining nine babies with RPR titers < four times higher than their mothers’ were managed as possible CS. All 12 babies with confirmed and possible CS were offered treatment with intravenous aqueous crystalline penicillin G (150,000 units/Kg/day) for 10 days, per local guidelines (16). Of these, five (41%) completed intravenous treatment, including two with confirmed CS; two babies were treated with oral azithromycin syrup at their mothers’ request as an alternative to intravenous treatment (17). Two babies initiated but did not complete IV treatment due to inability to remain at the health facility. The remaining babies did not initiate any form of treatment due to parental refusal. Repeat testing of babies and mothers was not performed due to resource constraints.

Follow-up calls to the six women whose babies could not be examined physically found that two infants were stillborn, one early neonatal death in a previously low-birth-weight baby (<2500g irrespective of gestational age) (21), and one additional low-birth-weight baby; the remaining two women could not be reached. Calls to the remaining high-risk women who were antibody positive but RPR non-reactive (whose babies were not examined or tested) (n=46) identified two stillbirths. Birth outcomes were not ascertained for the 47 high-risk women who tested negative for treponemal antibodies and were not tested using RPR, as these women were not followed up beyond antenatal screening.

## Discussion

This implementation study evaluated the service-delivery impact of a CFIR-guided, multi-component syphilis intervention across four public health facilities in Kampala, Uganda. The intervention combined same-day testing, on-site treatment, partner notification, and syphilis-exposed infant follow-up. Compared with the pre-intervention period, we found statistically significant increases in the number of women and partners tested for syphilis and in the number of those testing positive who received treatment. These gains occurred despite the COVID-19 pandemic, a major external disruption to health-system function (22), suggesting that the implementation strategy was robust enough to sustain service delivery under significant disruption. These findings are consistent with reports that reducing repeat clinic visits and transport costs improves uptake of antenatal care ANC syphilis services (23).

Partner notification and treatment improved substantially in relative terms, although it remained a matter of concern: even during implementation, more than a third of RDT-positive women’s partners were never tested. This mirrors our earlier Ugandan STOP trial, in which only 18.3% of male partners of pregnant women with syphilis received treatment (10).

The prevalence of treponemal antibody-positive syphilis among high-risk women (62%) was about fifteenfold higher than the general ANC population attending Kisenyi, Kawaala, and Kiswa (approximately 4%), consistent with previous research (24, 25). It is unclear if this was also confounded by lower health care access during COVID or systemic differences in the characteristics of pregnant women attending the two settings.

Follow-up of infants born to women with an RPR titer ≥1:8 revealed the most critical gap in the cascade: up to 46% met criteria for confirmed or possible CS (19) yet high treatment attrition was observed, with only five of twelve (41%) babies completing treatment, as opposed to the two babies who completed treatment with oral Azithromycin. The adverse outcomes observed among infants at risk of CS are consistent with previous research (26, 27).

Baseline gaps in screening and treatment likely reflect structural constraints like RDT/BPG stockouts and limited laboratory capacity, factors previously reported to reduce uptake of syphilis services (28, 29), underscoring the cross-contextual generalizability of these barriers. Project-supplied medications and diagnostics addressed stockout-related gaps, while task-sharing, increased staffing, and structured follow-up protocols improved follow-up of high-risk women and exposed infants where distance and costs limited in-person visits. Phone-based tracing was particularly feasible given that about 43% of Ugandans own a mobile phone (30).

This study has several strengths. It screened a very large number of women attending routine ANC, and used a pre- and during-intervention, multi-facility design with a CFIR-informed implementation strategy across lower-level ANC facilities and a tertiary referral hospital. This allowed systematic quantification of gains across the full maternal-partner-infant care cascade. Adjustment for site using a modified Poisson regression model strengthens the robustness of the reported risk differences. The project also established systematic follow-up of women with pregnancies at risk of CS and their infants, which was not routinely conducted before.

The study also has limitations. Because routine follow-up of high-risk women and their infants was not conducted before implementation, comparable pre-intervention data on pregnancy and infant outcomes were unavailable. CS was classified using the 2021 CDC guidelines adapted for low-resource settings, which may have increased the number of infants classified as possible CS, and treated infants were not retested. High-risk women were managed at a single referral site, limiting generalizability; maternal non-treponemal titers were unavailable; and women with discordant results (treponemal positive, RPR non-reactive) were excluded, potentially missing early or late infections. High-risk women who tested treponemal antibody-negative were not followed to birth, and the observational design without a concurrent control group precluded comparison of outcomes with lower-risk pregnancies or with pregnancies in which syphilis was present and untreated. RPR testing was conducted only at Kawempe with women at the other three sites treated on treponemal antibody positivity, which may have led to overtreatment and limits accurate estimation of the true syphilis burden. Finally, data collection relied on paper registers with manual tallying, which may have introduced transcription errors.

### Implications for policy and practice

Closing the partner- and infant-testing gaps requires structural rather than purely educational change. First, electronic data collection tools should be formally integrated into the Uganda EMR system to reduce reliance on manual processes and support timely decision-making. Second, a single, documented maternal-infant care pathway is needed, incorporating systematic flagging of exposed infants, synchronized appointments and dedicated tracing registers, so that follow-up does not depend on mothers recalling and reporting their diagnosis. Third, RPR titer testing is important to monitor serological cure or reinfections between pregnancies, guiding BPG use and preventing unnecessary repeat treatment. Sustaining improvements will depend on continued investment in reliable diagnostics and penicillin supply, task-shifted staff, and resources to facilitate post-partum follow-up of exposed infants. Affordable, rapid, lipoidal antibody-based infant diagnostics would reduce both overtreatment and missed cases of true CS.

## Conclusions

This CFIR-guided, multi-component implementation strategy substantially improved syphilis screening and same-day treatment coverage among pregnant women and their partners, even under pandemic constraints. However, significant gaps persist in testing and treating sexual partners and exposed infants. Closing these gaps will require education, strengthened supply chains, integrated same-day testing and treatment, enhanced health-worker capacity, and robust follow-up to improve maternal and newborn outcomes.

## Declarations

## Data Availability

The facility register data analyzed for this manuscript will not be made available online because of its sensitive nature, but the corresponding author can be contacted on a case-by-case basis to provide data following a reasonable request.

## Acknowledgements

The authors express their gratitude to all the study participants and the participating health facilities in Kampala District that allowed the study to be conducted within their premises. We are grateful to all the research assistants for their contribution to collecting data.

## Authors’ Contributions

All authors contributed to the study. AAO: Project administration, data collection, data analysis, draft manuscript writing. AAO is the guarantor; ABN: Project design and administration, supervision, manuscript reviews; ABM: Data collection, manuscript reviews; FD: Data management and analysis; YCM: Supervision, manuscript reviews; RK: Manuscript reviews; MMH: Supervision, manuscript reviews; RPR: Funding acquisition, project design, supervision, manuscript draft and reviews. All authors read and approved the final manuscript.

## Consent for publication

Not Applicable

## List of abbreviations

ANC: Antenatal Care
CDC: Centers for Disease Control
CFIR: Consolidated Framework for Implementation Research
CS: Congenital Syphilis
EMTCT: Elimination of mother-to-child transmission
HIV: Human Immunodeficiency Virus
IDI REC: Infectious Diseases Institute Research Ethics Committee
KSST: Kampala Syphilis Screening and Treatment project
PMTCT: Prevention of Mother-to-Child Transmission
RPR: Rapid Plasma Reagin
SAPN: Syphilis Assisted Partner Notification register
SBOR: Syphilis Birth Outcomes Register
SSA: Sub-Saharan Africa
TPHA: Treponema Pallidum Haemagglutination Assay
UDHS: Uganda Demographic and Health Survey
UNCST: Uganda National Council of Science and Technology
WHO: World Health Organization

## APPENDIX

