## Supplemental figure 1 for "Improving Antenatal Syphilis Screening and Treatment Coverage in Public Health Facilities in Kampala, Uganda: A Pre-Implementation and Implementation-Phase Study"

**Supplemental File 1 showing a map of Kampala city, showing divisions and health facilities where the study was conducted**

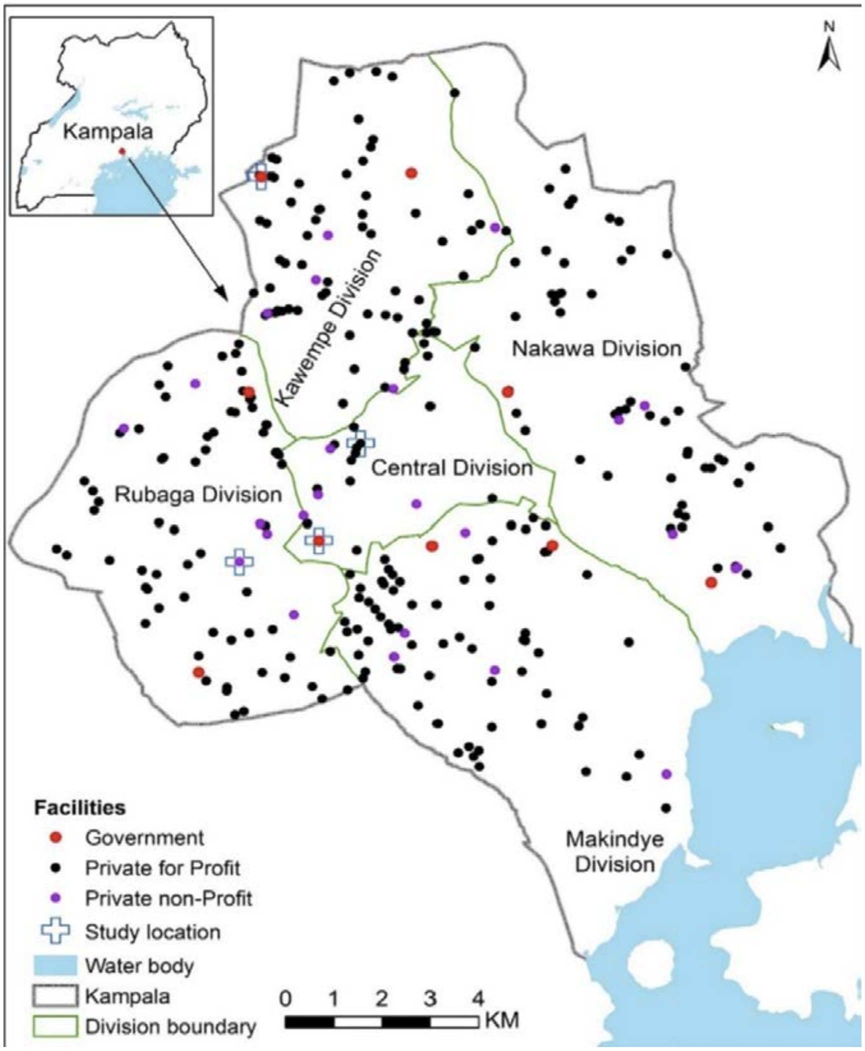


Map adopted from the Kampala Capital City Authority website. Available at [https://www.researchgate.net/figure/Map-of-Kampala-city-showing-divisions-and-health-facilities_fig1_383346122](https://www.researchgate.net/figure/Map-of-Kampala-city-showing-divisions-and-health-facilities_fig1_383346122)
