## Supplemental file 2 for "Improving Antenatal Syphilis Screening and Treatment Coverage in Public Health Facilities in Kampala, Uganda: A Pre-Implementation and Implementation-Phase Study"

### IDI- Syphilis APN Register Draft 1.0:
### SYPHILIS ASSISTED PARTNER NOTIFICATION (APN) REGISTER

NAME OF HEALTH FACILITY: ..........................................................CODE: .................LEVEL.........................

SUBCOUNTY/DIVISION: ..............................................................................................................................

HSD: ...........................................................................................................................................................

DISTRICT: ..........................................................................................................................................

DATE OPENED: ..........................................................DATE CLOSED: ..........................................................

INFECTIOUS DISEASES INSTITUTE
RESEARCH ETHICS COMMITTEE (IDI-REC)
APPROVED
VALID UNTIL
24 JUN 2023

### IDI SAPN Version 1.0: SYPHILIS ASSISTED PARTNER NOTIFICATION (SAPN) REGISTER

#### DESCRIPTION AND INSTRUCTIONS

To track all syphilis positive mothers that consent for APN and have at leastone sexual partner
**Copies:** One copy: This copy will be kept at the health facility unit
**Responsibility:** In charge of the MCH Clinic or APN focal person.

**PROCEDURE:**
1. Record all syphilis positive mothers eligible for APN and have consented for the service.
2. The DATE the register was started; NAME of health unit and the date the register was closed are written on the front cover. The register should be completed using data from the ANC client card, ANC Register and any additionalinformation as obtained from the respective client.
3. Pre-printed formats are available for this register, but in cases where they are not, counter books may be used for this register. If counter books are used, then draw lines and write headings, as shown in the Form below.
4. A serial number will be copied from the ANC register.
5. The health facility in-charge and DHO can determine additional data of local interest to monitor.

**KEY DEFINITIONS**
**Index client:** A syphilis positive mother eligible for APN
**Eligibility criteria for APN:** Newly identified syphilis positive.
**Partner/s:** An individual/s having or had sexual relationship with an index client in last 9 months.
**Notification:** The process of reaching out to or contacting the elicited partner/s of the index client.

**DESCRIPTION OF COLUMNS:**
The APN register should be confidential and thus the reason why it is not kept in the OPD.

**1: Client Initials and Serial Number:**
This is the first letter of the first name, middle name and surname e.g. Anne Mary Apio (AMA), the serial number should start with new Number, for every month.

**2: Upper row: Registration date:**
This is the date when an index client is recorded in this register

**2: Lower row: National ID# (NIN):**
National identification number as it appears on the client's national ID or Passport number.

**3: ANC serial Number:**
This denotes the ANC serial number as reflected on the ANC client card and ANC register.

**4: Date of Birth:**
This denotes the individuals date of birth. If individual cannot remember date of birth but knows the age, count back to get year of birth and use mid-month, mid-year to get the date of birth.

**5: Age in Years:**
This denotes the age in completed years.

**6. Sex (Female);**
Record sex of index client as Female

**7: Marital Status:** Record individual's marital status as; 1. Married,2. Cohabiting,3. Never married, 4. Separated, 5.Divorced, 6. Widow

**8: Telephone Contact:**
This denotes client's telephone contact(s) through which the client can be reached. Record the first telephone (1)
contact in the upper space and the alternative telephone contact in the lower space.

**9: Address:**
Record the physical address of the client (District, Sub-county, Parish, Village/Cell, Prominent feature)

**10: Index Client Type:**
Record the type of index client using any one of the codes below; 1. Newly identified (syphilis),
2. On ART with new risk (With syphilis), 3. Newly identified (HIV/syphilis), and 4. Referred from another ANC site

**11: Entry Point:**
Refers to the service point in the Health Care System where the client was identified or tested from. Record the appropriate entry point as;1. ANC, 2. Maternity

**12: SPECIAL CATEGORY**
Record client classification/ category as follows;
1. Prisoners
2. Refugees
3. Adolescent Girls and Young Women (AGYW)
4. People with Disabilities (PWD)
5. Others specify

**13: Number of Sexual Partners in the past 9 months:**
Record the number of sexual partners the index client has had in the past 9 months.

**14: Is this the first Syphilis APN encounter?**
Record if the client is eligible for APN for the first time or not by recording Yes/No

**15: Remarks:**
Record any additional comments important for client care that do not have a designated column

**16: Partner Name:**
This is the name of the partner, Upper row: Record surname, Lower row: Record first name

**17: Common Name:**
This denotes the name of the sexual partner which is commonly used/known by his community

**18: Upper row: Sex (Male);**
Record sex of sexual partner as Male, Lower row: Age in Years: This denotes the age in completed years.

**19: Upper row: Phone contact1:**
Record the first telephone contact of the partner, Lower row: Alternative Phone Contact: Record the alternative telephone contact of the partner.

INFECTIOUS DISEASES INSTITUTE
RESEARCH ETHICS COMMITTEE (IDI-REC)
APPROVED
VALID UNTIL
24 JUN 2023

### IDI SAPN Version 1.0: SYPHILIS ASSISTED PARTNER NOTIFICATION (APN) REGISTER

**20: Address:**
Record the physical address of the sexual Partner (District, Sub- County, Parish, Village/Cell, Prominent feature)

**21: Index client choice of notification:**
Denotes the choice of notification that the index client would prefer to use to reach this partner. Record code as: 1.Self with Call for Life Reminders (Date) and 2. Assisted/ Provider notification (Date)

**22: History of Intimate Partner Violence (IPV):**
Record Yes/No depending on whether there's any prior history of violence to the index client from this partner

**23: Notification Contact information:**
Contact Information: Denotes information about contacting the partner,
Upper row: First Contact Date: Denotes date of the first successful telephone contact to this partner. Middle row: Second Contact Date: Denotes date of the second successful telephone contact to this partnerLower row: Third Contact Date: Denotes date of third successful telephone contact to this partner
Means: Denotes the means of contacting the Sexual partner. Recorded using the codes below; 1. Phone. 2. ANC visit

Outcome: Record the follow-up outcome for each notification attempt using the codes below;
1. Came for Syphilis test
2. Will test elsewhere
3. Refused test
4. Already known Syphilis positive
5. Completed Syphilis treatment

**Codes for case closure:**
1. Partner has tested for Syphilis (Neg or Pos)
2. All attempts to locate partner have been unsuccessful over a period of 2 months,
3. Partner is confirmed to be already treated,
4. Partner is out of the district /country and confirmed will not be able to come for Syphilis testing at the health facility within the next 2 months,
5. Partner confirmed to be dead,
6. Partner is unwilling to come,
7. Partner has opted to test elsewhere,
8. Others specify

**24: Syphilis Testing:**
Denotes Syphilis Testing information for the sexual partner

**Tested for Syphilis:**
Denotes whether this partner has tested for Syphilis or not and the date tested for Syphilis.
**Upper Row:** Record Yes if partner has tested for Syphilis and No if has not tested for Syphilis (Yes/No)
**Lower Row:** Record date the sexual partner tested for Syphilis using the format dd/mm/yyyy

**Final Syphilis test Result:**
Denotes final Syphilis test result for partners that tested for Syphilis. Record according to the national Syphilis testing algorithm as;
Positive,
Negative, and
Inconclusive

**25: If Syphilis Positive:**
**Upper row;** Syphilis positive partners, record the code that applies to each using the following;
Newly identified to be treated and receive couple counselling
Already known Syphilis Positive received treatment
Already known Syphilis Positive not yet on treatment

**26: If Syphilis Negative, Linkage to:**
HIV negative partners, record the service to which the individual is linked using the codes below. More than one code may be used;
1. Couple counselling
2. Condom use
3. VMMC
4. Others (specify)

**27: Upper row –Was IPV experienced within1-month post notification:**
Determine if index client experienced GBV from this partner following notification and record presence of IPV as (Yes)
and absence of IPV as (No)
Lower row –Case closure status
Record codes as listed at the bottom of the register.

**28: Remarks:**
Record any additional comments important for client care that do not have a designated column.

INFECTIOUS DISEASES INSTITUTE
RESEARCH ETHICS COMMITTEE (IDI-REC)
APPROVED
VALID UNTIL
24 JUN 2023

### IDI SAPN Version 1.0: SYPHILIS ASSISTED PARTNER NOTIFICATION (APN) REGISTER

**INDEX CLIENT INFORMATION**

| 1 | 2 | 3 | 4 | 5 | 6 | 7 | 8 | 9 | 10 | 11 | 12 | 13 | 14 | 15 |
|---|---|---|---|---|---|---|---|---|---|---|---|---|---|---|
| Client ini | Registration date / National ID# (NIN): | Serial No | Date of Birth | Age in | Sex | Marital St | Telephone | Address *District* | Index Client Type *(Use Codes)* | Facility Entry | KP/PP Categorization: *(Use* | No. of Sexual Partners in past | Is this the first APN encounter? (Yes/No) | Remarks: |

**PARTNER INFORMATION**

| 16 | 17 | 1 | 19 | 20 | 2 | 22 | 23 | | | | 24 | 25 | 2 | 27 | 28 |
|---|---|---|---|---|---|---|---|---|---|---|---|---|---|---|---|
| Partner Name | CommonName / Age in Years | Sex | Telephone | Address | Index client choice of notification | History of Intimate Partner Violence (IPV) Any prior history of violence | Notification Contact information | | Tested for Syphilis (Yes/No) | | Syphilis Test | If Syphilis- | If Syphilis negative, Linked to | Was IPV experienced within 1-month post notification (Yes/No) | Remarks |
| | | | | | | | Means | Outcome | Date Tested | Case closure status (Use Codes) | Final Syphilis Tes | | | | |

| Col. 7. Marital Codes: | Col. 10 Index Client Type Codes: | Col. 11. Facility Entry Point Codes | Col. 12. Special Category (Codes) | Col 21. Index client choice of notification Codes: | Col. 23. Case closure codes: | Col 25 If Syphilis Positive code | Col 26. If Syphilis Negative code |
|---|---|---|---|---|---|---|---|
| 1=Married<br>2=Cohabiting<br>3=Never married<br>4=Separated<br>5=Divorced<br>6=Widowed | 1=Newly identified (syphilis)<br>2=On ART with new risk (With syphilis)<br>3=Newly identified (HIV/syphilis).<br>4. Referred from another ANC site | 1=ANC,<br>2=Maternity | 1= Prisoners<br>2=Refugees<br>3=Adolescent Girls and Young Women<br>4=People with Disabilities (PWD)<br>5=Others specify | 1=Self with call for life reminders (Date)<br>2= Assisted/Provider notification (Date) | 1=Partner has tested for Syphilis (Neg or Pos)<br>2=All attempts to locate partner have been unsuccessful over a period of 2 months,<br>3=Partner is confirmed to be already treated.<br>4=Partner is out of the district /country and confirmed will not be able to come for Syphilis testing at the health facility within the next 2 months.<br>5=Partner confirmed to be dead.<br>6=Partner is unwilling to come.<br>7=Partner has opted to test elsewhere.<br>8=Others specify | 1=Newly identified to be treated and receive couple counselling.<br>2=Already known Syphilis Positive received treatment<br>3=Already known Syphilis Positive not yet on treatment | 1=Couple counselling<br>2=Condom use<br>3=VMMC<br>4=Others (specify). |

INFECTIOUS DISEASES INSTITUTE RESEARCH ETHICS COMMITTEE (IDI-REC) APPROVED VALID UNTIL 24 JUN 2023

| | | | | | |
|---|---|---|---|---|---|
| | | | | **Col 23. Means Codes**<br>1=Phone<br>2= ANC visit | **Col 24 Outcome Codes**<br>1=Came for Syphilis test<br>2=Will test elsewhere<br>3=Refused test<br>4=Already known Syphilis positive<br>5=Completed Syphilis treatment |

INFECTIOUS DISEASES INSTITUTE
RESEARCH ETHICS COMMITTEE (IDI-REC)
APPROVED
VALID UNTIL
24 JUN 2023

### IDI- Syphilis APN Register Draft 1.0:
### SYPHILIS ASSISTED PARTNER NOTIFICATION (APN) REGISTER

NAME OF HEALTH FACILITY: ..........................................................CODE: ................LEVEL.......................

SUBCOUNTY/DIVISION: ..........................................................................................................................

HSD: ..............................................................................................................................................

DISTRICT: ......................................................................................................................................

DATE OPENED: ..........................................................DATE CLOSED: ..............................................................

INFECTIOUS DISEASES INSTITUTE
RESEARCH ETHICS COMMITTEE (IDI-REC)
APPROVED
VALID UNTIL
24 JUN 2023

### IDI SAPN Version 1.0: SYPHILIS ASSISTED PARTNER NOTIFICATION (SAPN) REGISTER

**DESCRIPTION AND INSTRUCTIONS**

To track all syphilis positive mothers that consent for APN and have at leastone sexual partner
**Copies:** One copy: This copy will be kept at the health facility unit
**Responsibility:** In charge of the MCH Clinic or APN focal person.

**PROCEDURE:**
1. Record all syphilis positive mothers eligible for APN and have consented for the service.
2. The DATE the register was started; NAME of health unit and the date the register was closed are written on the front cover. The register should be completed using data from the ANC client card, ANC Register and any additionalinformation as obtained from the respective client.
3. Pre-printed formats are available for this register, but in cases where they are not, counter books may be used for this register. If counter books are used, then draw lines and write headings, as shown in the Form below.
4. A serial number will be copied from the ANC register.
5. The health facility in-charge and DHO can determine additional data of local interest to monitor.

**KEY DEFINITIONS**
**Index client:** A syphilis positive mother eligible for APN
**Eligibility criteria for APN:** Newly identified syphilis positive.
**Partner/s:** An individual/s having or had sexual relationship with an index client in last 9 months.
**Notification:** The process of reaching out to or contacting the elicited partner/s of the index client.

**DESCRIPTION OF COLUMNS:**
The APN register should be confidential and thus the reason why it is not kept in the OPD.

**1: Client Initials and Serial Number:**
This is the first letter of the first name, middle name and surname e.g. Anne Mary Apio (AMA), the serial number should start with new Number. for every month.

**2: Upper row: Registration date:**
This is the date when an index client is recorded in this register

**2: Lower row: National ID# (NIN):**
National identification number as it appears on the client's national ID or Passport number.

**3: ANC serial Number:**
This denotes the ANC serial number as reflected on the ANC client card and ANC register.

**4: Date of Birth:**
This denotes the individuals date of birth. If individual cannot remember date of birth but knows the age, count back to get year of birth and use mid-month, mid-year to get the date of birth.

**5: Age in Years:**
This denotes the age in completed years.

**6. Sex (Female);**
Record sex of index client as Female

**7: Marital Status:** Record individual's marital status as; 1. Married,2. Cohabiting,3. Never married, 4. Separated, 5.Divorced, 6. Widow

**8: Telephone Contact:**
This denotes client's telephone contact(s) through which the client can be reached. Record the first telephone (1)
contact in the upper space and the alternative telephone contact in the lower space.

**9: Address:**
Record the physical address of the client (District, Sub-county, Parish, Village/Cell, Prominent feature)

**10: Index Client Type:**
Record the type of index client using any one of the codes below; 1. Newly identified (syphilis),
2. On ART with new risk (With syphilis), 3. Newly identified (HIV/syphilis), and 4. Referred from another ANC site

**11: Entry Point:**
Refers to the service point in the Health Care System where the client was identified or tested from. Record the appropriate entry point as;1. ANC, 2. Maternity

**12: SPECIAL CATEGORY**
Record client classification/ category as follows;
1. Prisoners
2. Refugees
3. Adolescent Girls and Young Women (AGYW)
4. People with Disabilities (PWD)
5. Others specify

**13: Number of Sexual Partners in the past 9 months:**
Record the number of sexual partners the index client has had in the past 9 months.

**14: Is this the first Syphilis APN encounter?**
Record if the client is eligible for APN for the first time or not by recording Yes/No

**15: Remarks:**
Record any additional comments important for client care that do not have a designated column

**16: Partner Name:**
This is the name of the partner, Upper row: Record surname, Lower row: Record first name

**17: Common Name:**
This denotes the name of the sexual partner which is commonly used/known by his community

**18: Upper row: Sex (Male);**
Record sex of sexual partner as Male, Lower row: Age in Years: This denotes the age in completed years.

**19: Upper row: Phone contact1:**
Record the first telephone contact of the partner, Lower row: Alternative Phone Contact: Record the alternative telephone contact of the partner.

INFECTIOUS DISEASES INSTITUTE
RESEARCH ETHICS COMMITTEE (IDI-REC)
APPROVED
VALID UNTIL
24 JUN 2023

### IDI SAPN Version 1.0: SYPHILIS ASSISTED PARTNER NOTIFICATION (APN) REGISTER

**20: Address:**
Record the physical address of the sexual Partner (District, Sub- County, Parish, Village/Cell, Prominent feature)

**21: Index client choice of notification:**
Denotes the choice of notification that the index client would prefer to use to reach this partner. Record code as; 1.Self with Call for Life Reminders (Date) and 2. Assisted/ Provider notification (Date)

**22: History of Intimate Partner Violence (IPV):**
Record Yes/No depending on whether there's any prior history of violence to the index client from this partner

**23: Notification Contact information:**
Contact Information: Denotes information about contacting the partner,
Upper row: First Contact Date: Denotes date of the first successful telephone contact to this partner. Middle row: Second Contact Date: Denotes date of the second successful telephone contact to this partnerLower row: Third Contact Date: Denotes date of third successful telephone contact to this partner
Means: Denotes the means of contacting the Sexual partner. Recorded using the codes below; 1. Phone, 2. ANC visit

Outcome: Record the follow-up outcome for each notification attempt using the codes below;
1. Came for Syphilis test
2. Will test elsewhere
3. Refused test
4. Already known Syphilis positive
5. Completed Syphilis treatment

**Codes for case closure:**
1. Partner has tested for Syphilis (Neg or Pos)
2. All attempts to locate partner have been unsuccessful over a period of 2 months,
3. Partner is confirmed to be already treated,
4. Partner is out of the district /country and confirmed will not be able to come for Syphilis testing at the health facility within the next 2 months,
5. Partner confirmed to be dead,
6. Partner is unwilling to come,
7. Partner has opted to test elsewhere,
8. Others specify

**24: Syphilis Testing:**
Denotes Syphilis Testing information for the sexual partner

**Tested for Syphilis:**
Denotes whether this partner has tested for Syphilis or not and the date tested for Syphilis.
**Upper Row:** Record Yes if partner has tested for Syphilis and No if has not tested for Syphilis (Yes/No)
**Lower Row:** Record date the sexual partner tested for Syphilis using the format dd/mm/yyyy

**Final Syphilis test Result:**
Denotes final Syphilis test result for partners that tested for Syphilis. Record according to the national Syphilis testing algorithm as;
Positive,
Negative, and
Inconclusive

**25: If Syphilis Positive:**
**Upper row;** Syphilis positive partners, record the code that applies to each using the following;
Newly identified to be treated and receive couple counselling
Already known Syphilis Positive received treatment
Already known Syphilis Positive not yet on treatment

**26: If Syphilis Negative, Linkage to:**
HIV negative partners, record the service to which the individual is linked using the codes below. More than one code may be used;
1. Couple counselling
2. Condom use
3. VMMC
4. Others (specify)

**27: Upper row –Was IPV experienced within1-month post notification:**
Determine if index client experienced GBV from this partner following notification and record presence of IPV as (Yes)
and absence of IPV as (No)
Lower row –Case closure status
Record codes as listed at the bottom of the register.

**28: Remarks:**
Record any additional comments important for client care that do not have a designated column.

INFECTIOUS DISEASES INSTITUTE
RESEARCH ETHICS COMMITTEE (IDI-REC)
APPROVED
VALID UNTIL
24 JUN 2023

### IDI SAPN Version 1.0: SYPHILIS ASSISTED PARTNER NOTIFICATION (APN) REGISTER

**INDEX CLIENT INFORMATION**

| 1 | 2 | 3 | 4 | 5 | 6 | 7 | 8 | 9 | 10 | 11 | 12 | 13 | 14 | 15 |
|---|---|---|---|---|---|---|---|---|---|---|---|---|---|---|
| Client Ini | Registration date / National ID# (NIN): | Serial No | Date of Birth | Age in | Sex | Marital | Telephone | Address *District* | Index Client Type *(Use Codes)* | Facility Entry | KP/PP Categorization: *(Use* | No. of Sexual Partners in past | Is this the first APN encounter? (Yes/No) | Remarks: |

**PARTNER INFORMATION**

| 16 | 17 | 18 | 19 | 20 | 21 | 22 | 23 | | | | 24 | 25 | 26 | 27 | 28 |
|---|---|---|---|---|---|---|---|---|---|---|---|---|---|---|---|
| | | | | | | | Notification Contact information | | | | | | | | |
| | | | | | | | | | Tested for Syphilis (Yes/No) | | | | | | |
| Partner Name | CommonName / Age in Years | Sex | Telephone | Address | Index client choice of notification | History of Intimate Partner Violence (IPV) Any prior history of violence | Means | Outcome | Date Tested | Case closure status (Use Codes) | Syphilis Test / Final Syphilis Test | If Syphilis | If Syphilis negative, Linked to | Was IPV experienced within 1-month post notification (Yes/No) | Remarks |

**Col. 7. Marital Codes:**
1=Married
2=Cohabiting
3=Never married
4=Separated
5=Divorced
6=Widowed

**Col. 10 Index Client Type Codes:**
1=Newly identified (syphilis)
2=On ART with new risk (With syphilis)
3=Newly identified (HIV/syphilis).
4. Referred from another ANC site

**Col. 11. Facility Entry Point Codes**
1=ANC,
2=Maternity

**Col. 12. Special Category (Codes)**
1= Prisoners
2=Refugees
3=Adolescent Girls and Young Women
4=People with Disabilities (PWD)
5=Others specify

**Col. 21. Index client choice of notification Codes:**
1=Self with call for life reminders (Date)
2= Assisted/Provider notification (Date)

**Col. 23. Case closure codes:**
1=Partner has tested for Syphilis (Neg or Pos)
2=All attempts to locate partner have been unsuccessful over a period of 2 months.
3=Partner is confirmed to be already treated.
4=Partner is out of the district /country and confirmed will not be able to come for Syphilis testing at the health facility within the next 2 months.
5=Partner confirmed to be dead.
6=Partner is unwilling to come.
7=Partner has opted to test elsewhere.
8=Others specify

**Col 25 If Syphilis Positive code**
1=Newly identified to be treated and receive couple counselling.
2=Already known Syphilis Positive received treatment
3=Already known Syphilis Positive not yet on treatment

**Col 26. If Syphilis Negative code**
1=Couple counselling
2=Condom use
3=VMMC
4=Others (specify).

INFECTIOUS DISEASES INSTITUTE RESEARCH ETHICS COMMITTEE (IDI-REC)
APPROVED
VALID UNTIL
24 JUN 2023

| | | | | | |
|---|---|---|---|---|---|
| | | | | **Col 23. Means Codes**<br>1=Phone<br>2= ANC visit | **Col 24 Outcome Codes**<br>1=Came for Syphilis test<br>2=Will test elsewhere<br>3=Refused test<br>4=Already known Syphilis positive<br>5=Completed Syphilis treatment |

INFECTIOUS DISEASES INSTITUTE
RESEARCH ETHICS COMMITTEE (IDI-REC)
APPROVED
VALID UNTIL
24 JUN 2023
