## Supplemental file 3 for "Improving Antenatal Syphilis Screening and Treatment Coverage in Public Health Facilities in Kampala, Uganda: A Pre-Implementation and Implementation-Phase Study"

### IDI-SBOR VERSION 1.0:

#### SYPHILIS BIRTH OUTCOME REGISTER

HEALTH UNIT NAME: ..........................................................................................................................CODE:............ LEVEL...

SUBCOUNTY/DIVISION:...........................................................................................................................................

HSD NAME.............................................................................................................................................................

DISTRICT:...............................................................................................................................................................

DATE OPENED: .................................................................................. DATE CLOSED:.................................................................................

INFECTIOUS DISEASES INSTITUTE
RESEARCH ETHICS COMMITTEE (IDI-REC)
APPROVED
VALID UNTIL
24 JUN 2023

DESCRIPTION AND INSTRUCTIONS

**Objective:** To record the deliveries and birth outcomes of syphilis positive pregnant mothers.

**Copies:** One copy and it stays at the health unit in the Maternity and post-natal ward

**Responsibility**: Personnel in-charge of the Maternity / post-natal Ward

**PROCEDURE:**

1. Write the name of the health unit, name of health sub district and district, the date the register opened and the date the register closed onthe front cover of the SBO Register.
2. Write all deliveries information (dates of delivery, names of the mother, Age, Parity, Admission Number, ANC number/REF IN Number /NB IPD number, address, baby diagnosis and treatment, Mother diagnosis and treatment) for all deliveries whether the mother delivers or not and also write the management procedures carried out and treatment given.
3. Counsel and test mothers who come to deliver with unknown syphilis status, and enter their Syphilis results in the appropriate column(column 16)
4. The SBO register is reviewed to document such things as: documenting the numbers of stillbirths, Non-immune hydrops, prematurity, small stature for gestational age, mucocutaneous signs, reticuloendothelial signs, and the medicines used to manage the patient.
5. All severe syphilis birth outcomes (stillbirth, Non-Immune hydrops, prematurity and small stature for gestational age) should be discussed in meetings.

**DESCRIPTION OF COLUMNS**

**ADMISSION INFORMATION:**

**(1) DATE / TIME OF ADMISSION (DOA):**
Write the date and time of admission-(day and month is sufficient for the date). It is best to use abbreviations for months (Jan, Feb. etc)

**(2) ADMISSION NO.** Write the serial number given to the mother receiving maternity services. This number begins with 1 at the beginning of each month

**(3) ANC NO / REF IN NO / C/REF IN:**
If the woman attended any Health Unit's ANC clinic, then theclient number on the ANC Card and the Health Unit's name (or abbreviation) is written. If the woman was referred (high risk delivery), write the REF IN and the REF IN No. in this column. For women referred from the Community, write C/REF IN. If woman did not attend ANC indicate NB for not booked anywhere.

**(4) IPD No:**
This is a unique serial number given to the mother during her stayin the Maternity Ward and begins with 1 at the beginning of the financial year (September).

**(5) NATIONAL IDENTIFICATION NUMBER (NIN):**
Write the National Identification Number for the client (If available).

**(6) CLIENT NAME:**
Enter both surname name and first name of the woman.

**(7) AGE:**
Enter the age of the woman in complete years

**(8) CLIENT CATEGORY:**
Write N – National, F – Foreigner and R – Refugee

**(9) ADDRESS:**
Enter the Client's phone number and address as indicated in the watermarks (District, sub-county, parish and village).

**(10) GRAVIDA:**
All the pregnancies the woman has carried should be entered in this column. For mothers admitted but did not deliver at that admission,note the gravida.

**(11) PARITY:**
This is the pregnancies carried beyond 7 months that the client hashad before (exclude the current pregnancy).

**(12) GESTATION AGE (WEEKS OF AMMENORRHEA):**
The age of the pregnancy in weeks is entered in this column, e.g. 34 to mean 34 completed weeks of gestation

**(13) TERM AND PRE TERM:**
Indicate if the delivery is term (T) or pre-term (P)
Term (T) refers to 37 completed weeks of Gestation and Pre-term isless than 37 weeks of Gestation

**(14) REASON FOR ADMISSION:**
Write the reason why the client was admitted such as in active labour,in false labour, Malaria in pregnancy, UTI, early rupture of membranes, Incomplete abortion etc

**DELIVERY INFORMATION:**

**(15) EMTCT-S CODE:**
Enter the eMTCT-S code for the woman (W) and her baby (B) that corresponds to the eMTCT-S services received. The codes used to record eMTCT-S services received by the client are listed below for both the initial and verification test.

1=Newly identified to be treated and receive counselling
2=Already known Syphilis Positive received treatment
3=Already known Syphilis Positive not yet on treatment

INFECTIOUS DISEASES INSTITUTE RESEARCH ETHICS COMMITTEE (IDI-REC) APPROVED VALID UNTIL 24 JUN 2023

**(16) RESULTS FOR SYPHILIS TESTS done at Maternity**

The screening results for Syphilis are to be documented in this column

Write the syphilis test results using the following codes:

- **Rx:** Client tested and results given – client reactive and given treatment
- **+Ve:** Client tested positive but not yet given treatment
- **NR:** Client tested results given- client non-reactive
- **NT:** Client not tested for syphilis

**(17) MODE OF DELIVERY**

The techniques used during delivery, such as, "Normal Vaginal delivery", "vacuum extraction ", "caesarean section", etc.

**(18) DATE OF DELIVERY:**

The date when the mother delivers is recorded in this column

**TIME OF DELIVERY:**

The time (in 12 hours) when the mother delivers is indicated in this column.If the delivery took place on the way to the facility, indicate BBA for Born Before Arrival

**CHILD INFORMATION:**

**(19) APGAR SCORE (A/S):**

Write the Apgar score both in the 1st and 5th minute in this column.

Add "A" if with Asphyxia e.g 5/10 A

**(20) SEX:**

Indicate the baby's sex. That is;

- M: For Male, and '
- F: For Female

**(21) REFERRED IN/FACILITY**

Please enter; 1. Referred in; Document name of the health facility

2. Facility

**(22) WEIGHT (WT):**

Indicate the weight of the baby in kilograms (e.g. 3.2 kgs)

**(23)** RISK STATUS:

Risk Status: Write "HIGH" for infants whose mothers: are identified late during pregnancy, labor and delivery, or postnatal period; have a syphilis positive test.Indicate "LOW" for infants whose mothers have received treatment for syphilis before third trimester pregnancy.

Depending on the infant's syphilis exposure status and the intervention to prevent mother-to-child transmission, write one of the following codes atthe duration in weeks where Benzathine penicillin was given:

- Yes: If Benzathine Penicillin was given in late 3rd trimester or has never been given.
- No: If Benzathine Penicillin/ any syphilis treatment was given earlier in pregnancy

**(24) TESTING & TREATMENT TO THE BABY:**

Write:

- Y: For "YES" if baby received SYPHILIS TEST and TREATMENT
- N: For "NO" if the baby has not received the test nor treatment (specify reasons).

1. Mother declined,
2. no test available,
3. lost to follow up
4. Others

INFECTIOUS DISEASES INSTITUTE
RESEARCH ETHICS COMMITTEE (IDI-REC)
APPROVED
VALID UNTIL
24 JUN 2023

**(25) TREATMENT OFFERED TO THE BABY:**

Write the treatment that was administered to the baby.

**(26) FINAL DIAGNOSIS FOR THE BABY- Syphilis associated signs found in the baby.**

Indicate the diagnosis the baby has at discharge

**Syphilis associated signs**

1. Stillbirth
2. Prematurity
3. Small stature for gestational age
4. Non-Immune hydrops
5. Congenital anemia
6. Snuffles (bloody mucoid nasal discharge)
6. None

**(27) CONDITION OF BABY AT DISCHARGE:**

Enter the condition of the baby at discharge as follows;

- MSB: Macerated still birth
- FSB: Fresh still birth
- NND: Immediate Neonatal Death (death within 24hrs)
- AL: Live Baby
- AL – R: Live baby referred to another unit.
- AL – T: Live baby transferred to another ward / department e.g Neonatal ward
- BDF: Baby born with defect

- (Please note that babies older than 1day are captured in postnatalregister)

**(28) COUNSELING AT DISCHARGE**

Enter C under Disch.if mother counseled on danger signs, home based care and when to return for PNC, enter NC under Disch.if not counseled on danger signs, home based care and when to return for PNC.

INFECTIOUS DISEASES INSTITUTE
RESEARCH ETHICS COMMITTEE (IDI-REC)
APPROVED
VALID UNTIL
24 JUN 2023

| 1 | 2 | 3 | 4 | 5 | 6 | 7 | 8 | 9 | 10 | 11 | 12 | 13 | 14 | 15 | 16 | | 17 | 18 | 19 |
|---|---|---|---|---|---|---|---|---|---|---|---|---|---|---|---|---|---|---|---|
| ADMISSION INFROMAION | | | | | | | | | | | | | | CHILD INFORMAION | | | | | |
| DOA / TOA | Admission No. | ANC No./Ref In No. / c/Ref In | IPD No. | NATIONAL IDENTIFICATION NUMBER (NIN) | Client Name | Age | Client Category | Address | Gravida | Parity | Gestation Age | Term(T)/ Pre-Term (P) | Reason For Admission | EMTCT-S CODE | RESULTS FOR SYPHILIS AND HIV TESTS | | MODE OF DELIVERY | DATE OF DELIVERY / TIME OF DELIVERY | APGAR SCORE (A/S) |
| | | | | | | | | | | | | | | | W | B | | | |

INFECTIOUS DISEASES INSTITUTE RESEARCH ETHICS COMMITTEE (IDI-REC)
APPROVED
VALID UNTIL
24 JUN 2023

| 20 | 21 | 22 | 23 | 24 | 25 | 26 | 27 | 28 |
|---|---|---|---|---|---|---|---|---|
| SEX | REFERRED IN/FACILITY | WEIGHT | RISK STATUS | TESTING & TREATMENT TO THE BABY | TREATMENT OFFERED TO THE BABY | FINAL DIAGNOSIS FOR THE BABY | CONDITION OF BABY AT DISCHARGE | COUNSELING AT DISCHARGE |

INFECTIOUS DISEASES INSTITUTE
RESEARCH ETHICS COMMITTEE (IDI-REC)
APPROVED
VALID UNTIL
24 JUN 2023

### IDI-SBOR VERSION 1.0:

#### SYPHILIS BIRTH OUTCOME REGISTER

**HEALTH UNIT NAME:** ........................................................................................................**CODE:**........... **LEVEL...**

**SUBCOUNTY/DIVISION:**....................................................................................................................................

**HSD NAME**..........................................................................................................................................

**DISTRICT:**...........................................................................................................................................

**DATE OPENED:** ................................................................... **DATE CLOSED:**................................................................

INFECTIOUS DISEASES INSTITUTE
RESEARCH ETHICS COMMITTEE (IDI-REC)
APPROVED
VALID UNTIL
24 JUN 2023

DESCRIPTION AND INSTRUCTIONS

**Objective:** To record the deliveries and birth outcomes of syphilis positive pregnant mothers.

**Copies:** One copy and it stays at the health unit in the Maternity and post-natal ward

**Responsibility**: Personnel in-charge of the Maternity / post-natal Ward

**PROCEDURE:**

1. Write the name of the health unit, name of health sub district and district, the date the register opened and the date the register closed onthe front cover of the SBO Register.
2. Write all deliveries information (dates of delivery, names of the mother, Age, Parity, Admission Number, ANC number/REF IN Number /NB IPD number, address, baby diagnosis and treatment, Mother diagnosis and treatment) for all deliveries whether the mother delivers or not and also write the management procedures carried out and treatment given.
3. Counsel and test mothers who come to deliver with unknown syphilis status, and enter their Syphilis results in the appropriate column(column 16)
4. The SBO register is reviewed to document such things as: documenting the numbers of stillbirths, Non-immune hydrops, prematurity, small stature for gestational age, mucocutaneous signs, reticuloendothelial signs, and the medicines used to manage the patient.
5. All severe syphilis birth outcomes (stillbirth, Non-Immune hydrops, prematurity and small stature for gestational age) should be discussed in meetings.

**DESCRIPTION OF COLUMNS**

**ADMISSION INFORMATION:**

**(1) DATE / TIME OF ADMISSION (DOA):**
Write the date and time of admission-(day and month is sufficient for the date). It is best to use abbreviations for months (Jan, Feb, etc)

**(2) ADMISSION NO.** Write the serial number given to the mother receiving maternity services. This number begins with 1 at the beginning of each month

**(3) ANC NO / REF IN NO / C/REF IN:**
If the woman attended any Health Unit's ANC clinic, then theclient number on the ANC Card and the Health Unit's name (or abbreviation) is written. If the woman was referred (high risk delivery), write the REF IN and the REF IN No. in this column. For women referred from the Community, write C/REF IN. If woman did not attend ANC indicate NB for not booked anywhere.

**(4) IPD No:**
This is a unique serial number given to the mother during her stayin the Maternity Ward and begins with 1 at the beginning of the financial year (September).

**(5) NATIONAL IDENTIFICATION NUMBER (NIN):**
Write the National Identification Number for the client (If available).

**(6) CLIENT NAME:**
Enter both surname name and first name of the woman.

**(7) AGE:**
Enter the age of the woman in complete years

**(8) CLIENT CATEGORY:**
Write N – National, F – Foreigner and R – Refugee

INFECTIOUS DISEASES INSTITUTE
RESEARCH ETHICS COMMITTEE (IDI-REC)
APPROVED
VALID UNTIL
24 JUN 2023

**(9) ADDRESS:**
Enter the Client's phone number and address as indicated in the watermarks (District, sub-county, parish and village).

**(10) GRAVIDA:**
All the pregnancies the woman has carried should be entered in this column. For mothers admitted but did not deliver at that admission,note the gravida

**(11) PARITY:**
This is the pregnancies carried beyond 7 months that the client hashad before (exclude the current pregnancy).

**(12) GESTATION AGE (WEEKS OF AMMENORRHEA):**
The age of the pregnancy in weeks is entered in this column, e.g. 34 to mean 34 completed weeks of gestation

**(13) TERM AND PRE TERM:**
Indicate if the delivery is term (T) or pre-term (P)
Term (T) refers to 37 completed weeks of Gestation and Pre-term isless than 37 weeks of Gestation

**(14) REASON FOR ADMISSION:**
Write the reason why the client was admitted such as in active labour,in false labour, Malaria in pregnancy, UTI, early rupture of membranes, Incomplete abortion etc

**DELIVERY INFORMATION:**

**(15) EMTCT-S CODE:**
Enter the eMTCT-S code for the woman (W) and her baby (B) that corresponds to the eMTCT-S services received. The codes used to record eMTCT-S services received by the client are listed below for both the initial and verification test.

1=Newly identified to be treated and receive counselling
2=Already known Syphilis Positive received treatment
3=Already known Syphilis Positive not yet on treatment

**(16) RESULTS FOR SYPHILIS TESTS done at Maternity**

The screening results for Syphilis are to be documented in this column

Write the syphilis test results using the following codes:

- **Rx:** Client tested and results given – client reactive and given treatment
- **+Ve:** Client tested positive but not yet given treatment
- **NR:** Client tested results given- client non-reactive
- **NT:** Client not tested for syphilis

**(17) MODE OF DELIVERY**

The techniques used during delivery, such as, "Normal Vaginal delivery", "vacuum extraction ", "caesarean section", etc.

**(18) DATE OF DELIVERY:**

The date when the mother delivers is recorded in this column

**TIME OF DELIVERY:**

The time (in 12 hours) when the mother delivers is indicated in this column.If the delivery took place on the way to the facility, indicate BBA for Born Before Arrival

**CHILD INFORMATION:**

**(19) APGAR SCORE (A/S):**

Write the Apgar score both in the 1st and 5th minute in this column.
Add "A" if with Asphyxia e.g 5/10 A

**(20) SEX:**

Indicate the baby's sex. That is;

- M: For Male, and '
- F: For Female

**(21) REFERRED IN/FACILITY**

Please enter; 1. Referred in; Document name of the health facility
2. Facility

**(22) WEIGHT (WT):**

Indicate the weight of the baby in kilograms (e.g. 3.2 kgs)

**(23)** RISK STATUS:

Risk Status: Write "HIGH" for infants whose mothers: are identified late during pregnancy, labor and delivery, or postnatal period; have a syphilis positive test. Indicate "LOW" for infants whose mothers have received treatment for syphilis before third trimester pregnancy.

Depending on the infant's syphilis exposure status and the intervention to prevent mother-to-child transmission, write one of the following codes adthe duration in weeks where Benzathine penicillin was given:

- Yes: If Benzathine Penicillin was given in late 3rd trimester or has never been given.
- No: If Benzathine Penicillin/ any syphilis treatment was given earlier in pregnancy

**(24) TESTING & TREATMENT TO THE BABY:**

Write:

- Y: For "YES" if baby received SYPHILIS TEST and TREATMENT
- N: For "NO" if the baby has not received the test nor treatment (specify reasons).

1. Mother declined,
2. no test available,
3. lost to follow up
4. Others

INFECTIOUS DISEASES INSTITUTE
RESEARCH ETHICS COMMITTEE (IDI-REC)
APPROVED
VALID UNTIL
24 JUN 2023

**(25) TREATMENT OFFERED TO THE BABY:**

Write the treatment that was administered to the baby.

**(26) FINAL DIAGNOSIS FOR THE BABY- Syphilis associated signs found in the baby.**

Indicate the diagnosis the baby has at discharge

**Syphilis associated signs**

1. Stillbirth
2. Prematurity
3. Small stature for gestational age
4. Non-Immune hydrops
5. Congenital anemia
6. Snuffles (bloody mucoid nasal discharge)
6. None

**(27) CONDITION OF BABY AT DISCHARGE:**

Enter the condition of the baby at discharge as follows;

- MSB: Macerated still birth
- FSB: Fresh still birth
- NND: Immediate Neonatal Death (death within 24hrs)
- AL: Live Baby
- AL – R: Live baby referred to another unit.
- AL – T: Live baby transferred to another ward / department e.g Neonatal ward
- BDF: Baby born with defect

- (Please note that babies older than 1day are captured in postnatalregister)

**(28) COUNSELING AT DISCHARGE**

Enter C under Disch.if mother counseled on danger signs, home based care and when to return for PNC, enter NC under Disch.if not counseled on danger signs, home based care and when to return for PNC.

INFECTIOUS DISEASES INSTITUTE
RESEARCH ETHICS COMMITTEE (IDI-REC)
APPROVED
VALID UNTIL
24 JUN 2023

| 1 | 2 | 3 | 4 | 5 | 6 | 7 | 8 | 9 | 10 | 11 | 12 | 13 | 14 | 15 | 16 | | 17 | 18 | | 19 |
|---|---|---|---|---|---|---|---|---|---|---|---|---|---|---|---|---|---|---|---|---|
| ADMISSION INFROMAION | | | | | | | | | | | | | | CHILD INFORMAION | | | | | | |
| DOA / TOA | Admission No. | ANC No./Ref In No. / c/Ref In | IPD No. | NATIONAL IDENTIFICATION NUMBER (NIN) | Client Name | Age | Client Category | Address | Gravida | Parity | Gestation Age | Term(T)/ Pre-Term (P) | Reason For Admission | EMTCT-S CODE | RESULTS FOR SYPHILIS AND HIV TESTS | | MODE OF DELIVERY | DATE OF DELIVERY | TIME OF DELIVERY | APGAR SCORE (A/S) |
| | | | | | | | | | | | | | | | W | B | | | | |

INFECTIOUS DISEASES INSTITUTE
RESEARCH ETHICS COMMITTEE (IDI-REC)
APPROVED
VALID UNTIL
24 JUN 2023

| | | | | | | | | | |
|---|---|---|---|---|---|---|---|---|---|
| | | | | | | | | SEX | 20 |
| | | | | | | | | REFERRED IN/FACILITY | 21 |
| | | | | | | | | WEIGHT | 22 |
| | | | | | | | | RISK STATUS | 23 |
| | | | | | | | | TESTING & TREATMENT TO THE BABY | 24 |
| | | | | | | | | TREATMENT OFFERED TO THE BABY | 25 |
| | | | | | | | | FINAL DIAGNOSIS FOR THE BABY | 26 |
| | | | | | | | | CONDITION OF BABY AT DISCHARGE | 27 |
| | | | | | | | | COUNSELING AT DISCHARGE | 28 |

INFECTIOUS DISEASES INSTITUTE
RESEARCH ETHICS COMMITTEE (IDI-REC)
APPROVED
VALID UNTIL
24 JUN 2023
