## Supplemental file 4 for "Improving Antenatal Syphilis Screening and Treatment Coverage in Public Health Facilities in Kampala, Uganda: A Pre-Implementation and Implementation-Phase Study"

**Supplemental File 4 shows the flow of study activities at all study sites during the implementation period**

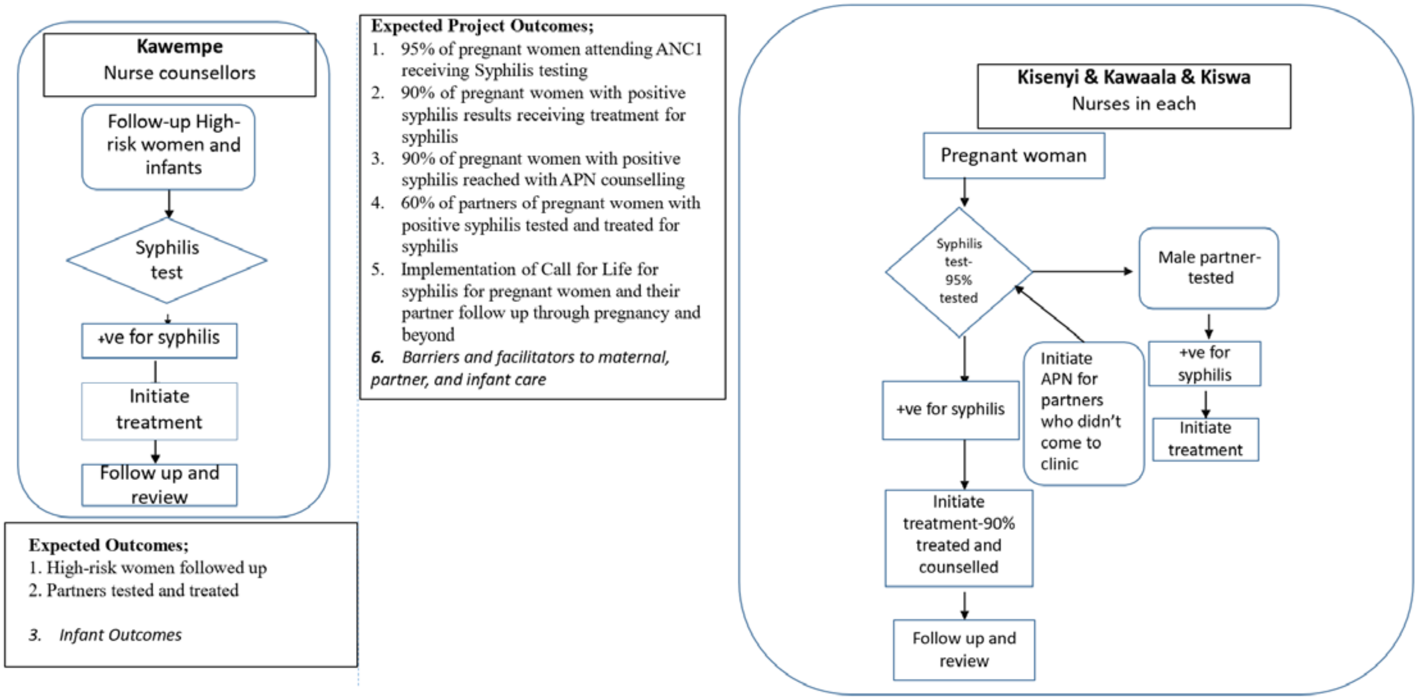
