## Supplemental File 5 for "Improving Antenatal Syphilis Screening and Treatment Coverage in Public Health Facilities in Kampala, Uganda: A Pre-Implementation and Implementation-Phase Study"

### Standards for Reporting Implementation Studies (StaRI)

Author checklist: 27 items, dual strands

| | |
|---|---|
| **Manuscript title** | **Improving Antenatal Syphilis Screening and Treatment Coverage in Public Health Facilities in Kampala, Uganda: A Pre-Implementation and Implementation-Phase Study** |
| **Corresponding author** | Rosalind Parkes-Ratanshi, PhD<br> |
| **Target journal** | Sexually Transmitted Infections: Topical Collection |
| **Study design (e.g. pre-post / before-and-during)** | Before-and-During Implementation |
| **Companion reporting guideline (e.g. TREND, SQUIRE 2.0, STROBE)** | STROBE |

**Note:** Item topics below are shortened summaries from the official item wording from Pinnock H et al. or the checklist on the EQUATOR Network

| # | Section | Item topic | Implementation strategy (column 1) | Intervention being implemented (column 2) | Page / section |
|---|---|---|---|---|---|
| **Title and abstract** | | | | | |
| **1** | Title | Identify the paper as an implementation study and state the methodology in the title and/or keywords | Improving Antenatal Syphilis Screening and Treatment Coverage in Public Health Facilities in Kampala, Uganda: A Pre-Implementation and Implementation-Phase Study | Syphilis testing and treatment | 1 |

| # | Section | Item topic | Implementation strategy (column 1) | Intervention being implemented (column 2) | Page / section |
|---|---|---|---|---|---|
| **2** | Abstract | Identify as an implementation study; describe the implementation strategy, the evidence-based intervention being implemented, and the key implementation and health outcomes | This is a pre-implementation and implementation-phase study; keywords include Implementation Research and Consolidated Framework for Implementation Research (CFIR)<br>The strategy was guided by CFIR, with same-day dual HIV/syphilis antibody testing, on-site treatment and partner notification at three ANC facilities, plus infant follow-up at a tertiary hospital. Facility registers data compared pre-implementation (Jan-Dec 2020) with implementation (Jun 2021-Jul 2022). | Same-day dual HIV/syphilis testing and treatment of women and partners, and follow-up of infants of high-risk women. Key results: screening 55.2% to 88.3%; same-day treatment 74% to 100%; partner testing 11.8% to 62%; 62% of high-risk women positive; up to 46% of examined infants had confirmed or possible CS, 41% of whom completed intravenous treatment. | Abstract |
| **Introduction** | | | | | |
| **3** | Introduction | Identify the problem: healthcare or public health deficiency the intervention aims to address | Implementation gap: syphilis screening and treatment coverage in Uganda remain below national and WHO targets (≥95% testing and treatment) despite the 2017 PMTCT guidelines, because of late ANC initiation, workforce shortages, long waits, and stockouts of RDTs and benzathine penicillin G. Few Ugandan studies have evaluated a framework-guided service-delivery intervention for this gap. | Health problem: untreated maternal syphilis causes stillbirth, abortion, prematurity and congenital syphilis (50-80% of cases). WHO estimated over 700,000 CS cases worldwide in 2022. The STOP trial in Kampala found 3.5% maternal syphilis prevalence, with untreated partners associated with adverse birth outcomes. | Introduction, paragraphs 1-4 |

| # | Section | Item topic | Implementation strategy (column 1) | Intervention being implemented (column 2) | Page / section |
|---|---|---|---|---|---|
| **4** | Introduction | Rationale for the implementation strategy and for the intervention | CFIR was used to systematically assess multi-level barriers and facilitators (with attention to inner-setting constraints) and to design strategies for improving rapid syphilis testing and treatment uptake. Barriers cited: late ANC initiation (only ~37% start in 1st/2nd trimester), workforce shortages, long waits, and RDT and benzathine penicillin G (BPG) stockouts. Needs assessment of 2020 register data identified service gaps during the pre-project needs assessment. | Same-day dual HIV/syphilis testing with immediate BPG treatment is in Uganda's national PMTCT guidelines (2017) and supports the WHO EMTCT targets (≥95% testing and ≥95% treatment). Partner treatment is needed to prevent reinfection of women which has previously been shown to be associated with adverse birth outcomes. | Introduction, paragraphs 2-4; Methods: Pre-project needs assessment |
| **5** | Aims and objectives | State aims, separating implementation objectives from intervention objectives | The Implementation objective was to evaluate whether a multi-component, CFIR-guided service-delivery intervention can close the gap between national targets and actual screening and treatment coverage, by comparing pre-implementation and implementation-period coverage across the mother-partner-infant cascade. | Intervention targets set from the needs assessment: syphilis screening of 95% of women at first ANC visit; treatment of 90% of those testing positive; partner-notification counselling for 90%; testing and treatment of 60% of partners. | Introduction, last paragraph; Methods: Pre-project needs assessment |
| **Methods** | | | | | |

| # | Section | Item topic | Implementation strategy (column 1) | Intervention being implemented (column 2) | Page / section |
|---|---|---|---|---|---|
| **6** | Methods | Study design and key features of the evaluation; cross-reference design-specific reporting standards; report protocol changes with reasons | Before-and-during implementation evaluation of routine facility register data, with no concurrent control group.<br>Pre-implementation: January-December 2020. Implementation: June 2021-July 2022. CFIR-guided. Four public facilities (three high-volume Health Centre IVs and one national referral hospital).<br>Risk differences from modified Poisson regression, adjusted for site.<br>No Design-specific reporting standards or protocol changes happened. | Quantitative service-delivery outcomes across the mother, partner and infant syphilis care cascade. | Abstract; Methods (Project setting, Data analysis); Discussion (Limitations) |

| # | Section | Item topic | Implementation strategy (column 1) | Intervention being implemented (column 2) | Page / section |
|---|---|---|---|---|---|
| **7** | Methods | Context in which the intervention was implemented (social, economic, policy, healthcare, organisational barriers and facilitators) | Kampala District, Uganda: urban public facilities.<br>Policy: same-day dual HIV/syphilis testing and treatment in national PMTCT guidelines since 2017, free in public facilities.<br>Barriers: late ANC initiation (~37% start in 1st/2nd trimester; 68% complete 4 visits), workforce shortages, long waits, stockouts, limited laboratory capacity, no confirmatory testing at lower-level sites (funding), COVID-19 lockdown.<br>Facilitators: high ANC attendance (99%), specialist capacity at Kawempe, ~43% mobile phone ownership. | Testing and treatment delivered within routine ANC, delivery and postnatal services, with project nurses and a Monitoring and Evaluation specialist at each site, project-supplied kits and BPG, and a hub-and-spoke laboratory for confirmatory RPR/TPHA. | Introduction, paragraph 3; Methods: Project setting; Discussion, paragraphs 1 and 5-6 |
| **8** | Methods | Targeted sites and populations | Sites: Kisenyi, Kawaala and Kiswa Health Centres IV, and Kawempe National Referral Women's Hospital.<br>Populations: at the three health centres, all pregnant women at their first ANC visit (<27 weeks) and male partners of women testing positive. At Kawempe, 'high-risk' women (never tested before 36 weeks, or testing positive in the third trimester, at delivery or postnatally), their partners and their infants. | Intervention recipients: pregnant women (RDT screening and treatment), partners of positive women (testing and treatment), and infants born to women with RPR titre ≥1:8 (examination, RPR testing, treatment). | Methods: Project setting; Project population |

| # | Section | Item topic | Implementation strategy (column 1) | Intervention being implemented (column 2) | Page / section |
|---|---|---|---|---|---|
| **9** | Methods | Description of the implementation strategy and of the intervention | CFIR-guided pre-implementation needs assessment of 2020 register data against Ministry of Health readiness tools, which set targets and led to new data tools (electronic Syphilis APN register and the Syphilis Birth Outcomes Register).<br>Project nurses and an M&E specialist at each site; sensitisation sessions on syphilis; referral of at-risk women from the three health centres to Kawempe; project-supplied kits and BPG; monthly register reviews; phone follow-up after delivery. | SD Bioline HIV/syphilis Duo rapid test, with same-day BPG for positives (return for 2 further weekly doses) and partner-notification counselling. Partners tested, with a single BPG dose if positive.<br>At Kawempe: confirmatory RPR/TPHA, RPR titres (high ≥1:8), infants examined and tested, CS classified per adapted CDC criteria, and 10-day IV aqueous crystalline penicillin G (150,000 units/kg/day) for confirmed or possible CS. | Methods: Pre-project needs assessment; Project implementation |
| **10** | Methods | Subgroups recruited for additional research tasks and/or nested studies | Not Done | Not Done | N/A |

| # | Section | Item topic | Implementation strategy (column 1) | Intervention being implemented (column 2) | Page / section |
|---|---|---|---|---|---|
| **11** | Methods | Outcomes (implementation outcomes and health outcomes) | Service-delivery outcomes, with targets: screening of 95% of first-ANC attendees; treatment of 90% of positives; partner-notification counselling for 90% of positive women; testing and treatment of 60% of partners.<br>Partners testing positive and treated; treatment completion (3 BPG doses)<br>Infant follow-up, testing and IV treatment completion at Kawempe. | Health outcomes (descriptive, Kawempe only): syphilis positivity in women and partners; RPR reactivity and titres; infant RPR reactivity and CS classification (confirmed or possible); stillbirth, low birth weight and early neonatal death. | Methods: Pre-project needs assessment, Project implementation |
| **12** | Methods | Process evaluation: objectives and outcomes related to how the strategy is expected to work | Formal process evaluation of how the strategy was expected to work was done using CFIR.<br>Monthly register reviews at each site tracked service-delivery outcomes (proportion of women and partners screened, tested and treated; CFIR was used for the needs assessment and process evaluation. | Monthly register reviews at each site tracked service-delivery outcomes (proportion of women and partners screened, tested and treated; CFIR was used for the needs assessment and process evaluation. | Methods: Project implementation<br>CFIR figure |
| **13** | Methods | Economic evaluation | No economic evaluation was done | No economic evaluation was done | N/A |

| # | Section | Item topic | Implementation strategy (column 1) | Intervention being implemented (column 2) | Page / section |
|---|---|---|---|---|---|
| **14** | Methods | Rationale for sample size (calculation, budgetary or practical constraints, data saturation, as appropriate) | No sample size calculation reported. All eligible women attending during the study period were included (all first-ANC attendees at the three health centres; all identified high-risk women at Kawempe). | N/A | N/A |
| **15** | Methods | Methods of analysis, with reasons for the choice | Data were manually extracted from the registers at all sites and tallied as daily counts as the electronic medical records system was still being set up across the country in all public health facilities.<br><br>Proportions were calculated; pre-implementation and implementation outcomes compared as proportions and differences.<br><br>Risk differences and 95% CIs from a modified Poisson regression with robust standard errors, adjusted for site. Reasons for choosing this model are not stated. | Kawempe maternal, and infant outcomes are reported descriptively as counts and proportions, with no pre-post comparison or statistical testing. | Methods: Data analysis |
| **16** | Methods | A priori subgroup analyses (for example by site or population) | None Done. The analysis was adjusted for site but not stratified by site. Kawempe high-risk women were analysed as a separate group, not as a pre-specified subgroup. | None Done. | N/A |
| **Results** | | | | | |

| # | Section | Item topic | Implementation strategy (column 1) | Intervention being implemented (column 2) | Page / section |
|---|---|---|---|---|---|
| **17** | Results | Populations | Lower-level facilities (Kisenyi, Kawaala, Kiswa): 33,831 women attended a first ANC visit in 2020 (pre-implementation); 12,611 first-ANC attendees were sensitised during implementation, 11,133 tested.<br>Partners of positive women were also included.<br>Kawempe National Referral Hospital: 125 high-risk women identified during implementation (47 treponemal-negative, 78 positive). | Pre-implementation: 18,684 screened; 740 positives; 547 treated; 87 partners tested.<br>Implementation: 11,133 tested; 335 positives (all treated); 208 partners tested; 49 partner positives.<br>Kawempe: 78 positive women; 54 partners tested; 32 RPR-reactive; 26 of 32 infants followed up (Figures 3-4). | Results: all sub-sections; Figures 3-4 |
| **18** | Results | Outcomes | Screening: 55.2% to 88.3% (+33.63; 95% CI 20.99-46.27; p<0.001; Table 2).<br>Same-day treatment: 74% to 100% (+26.11; 22.37-29.86; p<0.001).<br>Partner-notification counselling: 100% (335/335). Partner testing: 11.8% to 62% (+51.91; 46.78-57.04; p<0.001).<br>Partner treatment: 100% of positives (difference 29.60; 19.95-39.25; p<0.001).<br>Targets met for treatment, counselling and partner testing; 95% screening target not reached (88.3%). | Kawempe high-risk women: 62% (78/125) syphilis-positive; 41% (32/78) RPR-reactive, 90.6% of these titre ≥1:8; 55% (43/78) completed 3 BPG doses. Infants: 26/32 followed up; 12 (46%) RPR-reactive; 3 confirmed and 9 possible CS; 5/12 (41%) completed IV treatment.<br>Adverse outcomes: 4 stillbirths, 1 early neonatal death, 2 low-birth-weight babies.<br>No pre-implementation comparison for infant outcomes was done since this was not being implemented, hence the scarcity of data. | Results: all sub-sections; Table 2; Figures 3-4 |

| # | Section | Item topic | Implementation strategy (column 1) | Intervention being implemented (column 2) | Page / section |
|---|---|---|---|---|---|
| **19** | Results | Process evaluation data mapped to the mechanism by which the strategy is expected to work | Process-evaluation data were mapped to CFIR constructs or to the mechanism of effect. The Discussion also offers possible explanations (dedicated staffing, project-supplied kits and BPG, same-day service, task-sharing, phone tracing), but these were not tested. | N/A | Discussion, paragraphs 1 and 5<br>CFIR constructs figure |
| **20** | Results | Economic evaluation | Not done | Not done | N/A |
| **21** | Results | Representativeness and outcomes of subgroups, including nested research tasks | No nested studies and no priori subgroup analyses were conducted.<br>Kawempe high-risk women were described as a separate group, analysed descriptively with no pre-implementation comparison. Representativeness was hence limited since high-risk women were managed at a single referral site. | Outcomes for the 125 high-risk women and their infants (see item 18).<br>The 47 treponemal-negative high-risk women were not followed for birth outcomes.<br>Birth outcomes were not ascertained for 2 women who could not be reached. | Results: Kawempe and Infant outcomes; Discussion (Limitations) |

| # | Section | Item topic | Implementation strategy (column 1) | Intervention being implemented (column 2) | Page / section |
|---|---|---|---|---|---|
| **22** | Results | Fidelity and adaptation | Strategies aimed to improve fidelity to national guidelines; women and partners were treated per Ugandan guidelines.<br>Adaptations: no confirmatory testing at the three lower-level sites (due to funding limitations); women unable to return were given later BPG doses and told to finish at the nearest facility; phone follow-up; paper-based registers (electronic versions not integrated into the EMR). | Kawempe: 55% (43/78) completed all 3 BPG doses; completion not ascertained for the remainder. CDC Congenital syphilis case definition adapted for low-resource settings (no CSF or long-bone radiographs).<br>Two infants received oral azithromycin at maternal request; repeat testing not done (resource constraints).<br>Three-dose completion at lower-level sites not reported. | Methods: Needs assessment, Project implementation; Results: Kawempe; Discussion (Limitations) |
| **23** | Results | Contextual changes, if any, that may have affected outcomes | The implementation period coincided with the COVID-19 lockdown in Uganda, with movement restrictions and transport disruptions affecting service delivery and ANC attendance.<br>Project-supplied test kits and medication addressed stockouts.<br>No concurrent control group to separate these effects. | Possible confounding of the high prevalence in Kawempe high-risk women by reduced healthcare access during COVID-19 or systemic differences between the two settings | Methods: Project implementation; Discussion, paragraphs 1 and 3 |

| # | Section | Item topic | Implementation strategy (column 1) | Intervention being implemented (column 2) | Page / section |
|---|---|---|---|---|---|
| **24** | Results | All-important harms or unintended effects in each group | No harm or unintended effects occurred. There was possible overtreatment of women with previously treated syphilis or false-positive RDTs, because the three lower-level sites treated on RDT result alone (Discussion: Limitations). | Adverse pregnancy and infant outcomes (stillbirths, low birth weight, early neonatal death) are reported as outcomes in at-risk pairs and not attributed to the intervention. | Results: Infant outcomes; Discussion (Limitations) |
| **Discussion** | | | | | |
| **25** | Discussion | Summary of findings, strengths and limitations, comparison with other studies, conclusions | Summary: coverage of testing and same-day treatment improved significantly, despite COVID-19; partner testing rose nearly five-fold but over a third of partners remained untested.<br>Strengths: large numbers screened, multi-facility design, CFIR-informed strategy, site-adjusted analysis.<br>Limitations: pre-post design with no control group, no pre-intervention infant outcomes, adapted CS case definition, single referral site, RDT-only treatment at lower-level sites, paper-based data.<br>Comparison with previous studies was done | High syphilis prevalence in high-risk women (62%) compared with the general ANC population (about 4%); major gap in infant treatment completion (5 of 12).<br>Conclusions: The strategy improved screening and treatment coverage; partner and infant gaps persist. | Discussion, paragraphs 1-6; Conclusions |

| # | Section | Item topic | Implementation strategy (column 1) | Intervention being implemented (column 2) | Page / section |
|---|---|---|---|---|---|
| **26** | Discussion | Implications | Structural rather than educational changes are urgently needed: integrate electronic data tools into the Uganda EMR; a single maternal-infant care pathway (flagging of exposed infants, synchronised appointments, tracing registers); sustained investment in diagnostics, BPG supply, dedicated staff or task-shifting, and airtime for follow-up. | RPR titre testing to monitor serological cure or reinfection; affordable rapid lipoidal infant diagnostics.<br>Conclusions: parental education, stronger supply chains, earlier ANC attendance, integrated same-day testing and treatment, phone-based follow-up (Kampala and similar settings). | Discussion (final paragraph); Conclusions |
| **General** | | | | | |
| **27** | General | Regulatory approvals (ethics, use of routine data, governance), study registration or protocol availability, funding, conflicts of interest | Ethics: Infectious Diseases Institute REC (IDI REC 008/2022) and Uganda National Council for Science and Technology (HS2436ES). Informed consent waived for this quantitative sub-study (routine service-delivery data collected as standard of care).<br>The authors have no conflict of interest to declare.<br>Funding declaration stated in acknowledgements section. | As column 1. | Methods: Ethical approval<br>Acknowledgements |

Pinnock H, Barwick M, Carpenter CR, Eldridge S, Grandes G, Griffiths CJ, et al. Standards for Reporting Implementation Studies (StaRI): explanation and elaboration document. BMJ Open. 2017;7(4):e013318. doi:10.1136/bmjopen-2016-013318
